# Characterising the depression pathway in secondary care: a UK-based epidemiological study of patient characteristics, comorbidities, and treatments

**DOI:** 10.1101/2025.07.15.25331272

**Authors:** Laura A. Hannah, Laura Angco, Emanuele F. Osimo, Jonathan R. Lewis, Cathy M. Walsh, Rudolf N. Cardinal

## Abstract

**BACKGROUND:** Depression is a disabling disorder with variable outcomes. In severe cases treatment is provided by specialist mental health care services, yet there is a lack of real-world evidence demonstrating how depression is managed within these settings, and consequently, a limited understanding of how to improve care for this population.

**AIMS:** We examine the characteristics of patients receiving secondary mental healthcare for depressive disorders within a UK National Health Service (NHS) provider, and the treatments they receive. We investigate when patients receive treatments, and what predicts the use of specific treatments, improvement, and duration with services, with the aim of comparing real-world care to that advised by national guidelines.

**METHODS:** A retrospective cohort study was conducted using de-identified electronic patient records of patients with depression referred to Cambridgeshire and Peterborough NHS Foundation Trust (serving a population ∼0·86 million), between January 2013 and June 2021. ANOVA models examined predictor variables of improvement and duration of care, while survival analyses explored treatment initiation rates and predictors of which treatments were used.

**RESULTS:** 9,083 patients met the study’s inclusion criteria. Almost half of those with depression had additional psychiatric diagnoses, reflecting the complexity of cases in secondary care. Treatment within secondary care was associated with improvements in both depressive and overall symptoms. Patients with a greater degree of psychiatric co-morbidity and those with lower socio-economic status indicators presented with greater overall illness severity at baseline, were more likely to be admitted into hospital, spent longer with services, and improved less than the average. Treatment patterns differed across age groups, sex/gender, socio-economic status, and psychiatric comorbidities. Some nationally recommended further-line treatments appeared to be under-used.

**CONCLUSIONS:** Treatment gaps in further-line treatments for depression exist, highlighting key areas for service improvement. Future work should target patients with complex needs and those who are socio-economically deprived.

## 1. INTRODUCTION

Depression is a debilitating disorder with variable life course affecting 280 million people worldwide (1). In the United Kingdom (UK), an estimated one in six adults experience moderate to severe depressive symptoms (2). Despite receiving multiple lines of treatments, approximately a third of patients suffer persistent depressive symptoms (3), with the most severe requiring support from specialist services. UK guidelines provide further-line treatment recommendations, but a lack of real-world evidence demonstrating how depression and persistent depression is currently managed across secondary mental health (MH) settings hampers the improvement of treatment pathways.

### 1.1 Consequences and management of persistent depression

The term “persistent depression” is used to describe either prolonged depressive symptoms or treatment-resistant depression (TRD), itself most widely defined as failure to respond to two or more treatments at an adequate dose and duration (4). Persistent depression is associated with poor outcomes and high economic costs. Patients who do not respond to first- or second-line treatments are at increased risk of relapse and struggle to achieve remission (3,5), with as few as 13% sustaining remission after five years (6). These associations become even stronger with increasing numbers of treatment types and greater illness severity (3). Furthermore, many patients report suicidal ideation and/or suicide attempts (7). Alongside the personal distress, persistent depression is linked to greater resource use and productivity loss, imposing a heavy economic burden on healthcare systems and society (8).

When first-line psychological and/or pharmaceutical antidepressant treatments are ineffective, UK guidelines (9,10) advise either switching, increasing, or combining treatment. Should symptoms persist, or a patient is considered high risk, a referral to specialist services is recommended, where further line treatments should be considered, such as augmentation with second-generation antipsychotics (SGA), lithium, and/or electroconvulsive therapy (ECT) (9). However, recent studies indicate inconsistent adherence to these guidelines. Multiple studies report limited switching or combination treatment (11,12), and only 35% of TRD patients receive augmentative treatment (6). Furthermore, use of monoamine oxidase inhibitors (MAOIs), a further-line treatment, has substantially decreased (13,14). Persistent depression is an international problem, and while guidelines may differ, studies across Europe and the United States report similar outcomes and treatment gaps (15,16).

### 1.2 Gaps in evidence

There is a need for real-world evidence demonstrating how depression is managed, and treatment outcomes, in specialist MH care settings. Existing studies have predominantly relied on primary care data or trial outputs; however, data pertaining to those who experience the greatest illness severity is scarce. Where treatment in secondary care has been analysed, it has been hindered by incomplete prescription and diagnostic information, absence or minimal data relating to specialist treatments and/or outcome measures, recall bias, and small sample sizes (6,12,17). Given the specialist support required to tackle persistent depression, a comprehensive understanding of the management of depression within these services is necessary to improve outcomes; this has been recognised as a national research priority (18).

We aimed to use a large real-world secondary care dataset to examine the characteristics of those using secondary care services for depression, and the treatments they receive. We investigated when patients received treatments, and what variables predicted the use of specific treatments, improvement, and duration with services, with the aim of comparing real-word care to that advised by national guidelines.

The study did not employ an explicit definition of persistent depression, but included all patients in contact with the secondary care service with a diagnosis of a depressive disorder; by virtue of examining a secondary care service, most such patients are likely to have received multiple unsuccessful lines of treatment previously. A small proportion may present to specialist services without having previously received multiple treatments, such as first presentations of depressive illnesses to liaison psychiatry services, or severe/high-risk presentations requiring prompt secondary care input.

## 2. METHODS

### 2.1 Study design and setting

Retrospective cohort study using routinely collected UK National Health Service (NHS) patient data in a secondary MH care setting, Cambridgeshire and Peterborough NHS Foundation Trust (CPFT). **Figure 1** shows the study’s design, encompassing inclusion criteria and data collection time frames.

**Figure 1.**
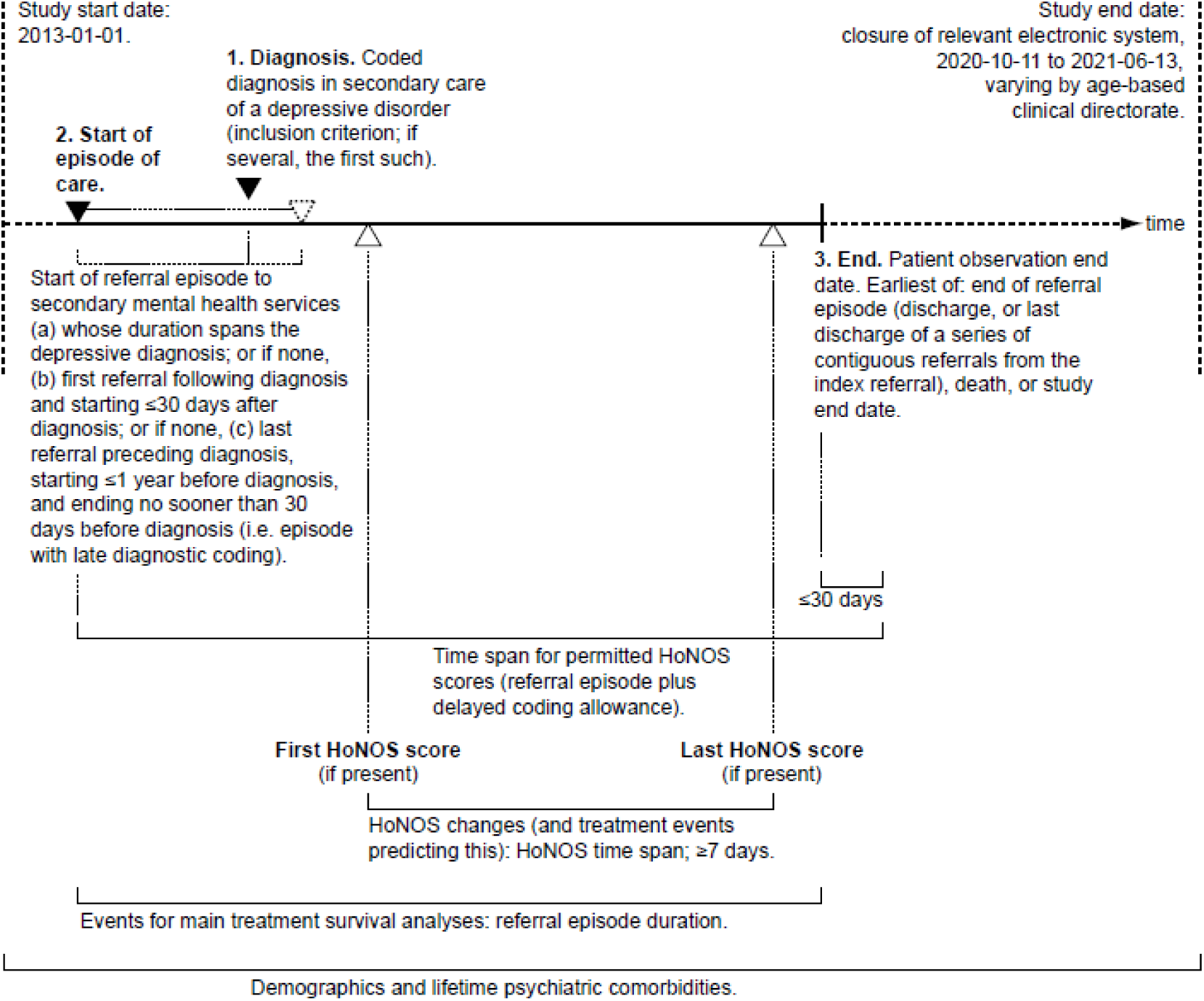
Timeline for each patient analysed in the study. Step 1: within the overall study start/end dates, patients were initially identified within CPFT data via a coded diagnosis representing a depressive disorder (see Methods), as shown. Step 2: The start of an episode of care associated with that diagnosis was identified, as shown. Step 3: The end of that episode of care was then identified. Outcome measures (HoNOS scores) were then extracted from within that episode of care, if present. (HoNOS, Health of the Nation Outcome Scales.)

Patient and public involvement, including individuals with lived experience of persistent depression, was incorporated into the development of the wider Informing VALues-based practice in Depression (i-VALiD) project of which this study is part of.

### 2.2 Data sources and ethical approval

The authors assert that all procedures contributing to this work comply with the ethical standards of the relevant national and institutional committees on human experimentation and with the Helsinki Declaration of 1975, as revised in 2013.

CPFT provides specialist secondary MH care to a catchment area of ∼0.86 million people. Its electronic patient records (EPRs) were de-identified into the CPFT Research Database; data from January 2013 to June 2021 were extracted for the purpose of this study. For approval details, see **Supplementary Methods.**

### 2.3 Patient selection and episode of care definition

Eligible patients included those referred to CPFT with a diagnosis, coded by a CPFT clinician according to the International Classification of Diseases (ICD-10) (19), of F32 (depressive episode) or F33 (recurrent depressive disorder), or the following diagnostic categories also involving the syndrome of depression: F20.4; F25.1; F31.3; F31.4; F31.5; F38.1; and F41.2 (see **Supplementary Methods** for full details).

Episodes of care were identified using dates pertaining to referral, depression diagnosis, and discharge (**Figure 1; Supplementary Methods**).

### 2.4 Data collection

All data were obtained from structured fields, except for medications (see below).

#### Demographic variables

We examined the following sociodemographic variables: sex (male/female), age at episode start (years), ethnicity (White/Asian/Black/Mixed/Other), and socio-economic status. Socio-economic status was measured using the standard England Index of Multiple Deprivation (IMD), based on a de-identified (blurred) version of the patient’s residential postcode. Patients were grouped into age categories corresponding to CPFT’s clinical Directorates, according to age at referral (see **Supplementary Methods** for all demographic measurement details).

#### Clinical and service activity data

Depressive diagnoses and lifetime other psychiatric morbidities (henceforth referred to as “comorbidity” without implied primacy) were obtained through diagnostic fields and categorised by ICD-10 sub-chapter (e.g. F1, F2). We also report the first recorded psychiatric diagnosis other than depression, and the timing of this in relation to the depression diagnosis.

Mortality data was available, via automatic linkage to national NHS Spine data, for those who died during the study timeframe.

Detentions under the UK Mental Health Act (MHA) (20) after referral were extracted. Treatment with ECT was identified via referral to a relevant service. Input from a psychologist in secondary care services was identified via a structured attribute/type of progress notes. See **Supplementary Methods** for other variables.

#### Medications

A previously validated natural language processing software tool (21) was used to extract mentions of medications from de-identified free text, prior to extraction of structured data for this study. We examined drugs recommended for depressive disorders by the UK National Institute for Health and Care Excellence (NICE) (9) and the British Association for Psychopharmacology (BAP) (22) (see **Supplementary Methods** for full list).

### 2.5 Outcome variables

#### Health of the Nation Outcome Scales (HoNOS)

The HoNOS, a clinician-rated 12-item instrument, measures patients’ behaviour, impairment, symptoms, and social functioning using a 5-point system (from 0 = no problem to 4 = severe-to-very-severe problem) (23). We examined depression sub-scores and total (overall) HoNOS scores. We took the first HoNOS measurement to reflect initial severity. Changes were only calculated for patients having two HoNOS reports at least seven days apart; changes were calculated as the last recorded score minus the first, with negative changes indicating improvement (reduction in symptoms). Missing data were not imputed.

#### Duration with secondary care services

Within secondary care, patients may receive treatment from multiple services. Such referrals were amalgamated into episodes of care, allowing for up to one day between services. For analysis of total duration with services, the durations were log-transformed in light of positive skew.

### 2.6 Statistical analyses

Categorical variables are reported as number (percentage) and continuous variables as mean (standard deviation). Chi-square contingency tests were used to compare demographics across the three directorates or versus the local population (24).

We analysed outcome scores and duration with services using linear models (analysis of variance [ANOVA]), with type III sums of squares (testing the effect of each predictor variable over and above all others). We show specific contrast tests where an ANOVA term was significant. A sensitivity analysis was performed by excluding those with a lifetime diagnosis of bipolar affective disorder (BPAD), given the likely changes in treatment that this diagnosis might entail.

We used survival analyses to determine the proportion of patients initiating nationally (NICE/BAP) recommended treatments, as well as the timing of these treatment initiations, during the course of care in secondary MH services. Data were censored for death, discharge from secondary care services, and the date at which the Servelec RiO EPR system was last used for each clinical Directorate (CAMH 2020-10-11; Adult 2020-12-06; Older Adult 2021-06-13). The event of interest was defined as the date of first mention of a given treatment. Patients were therefore excluded from that analysis if they were referred to and discharged from CPFT on the same day, but were also excluded if that treatment had occurred prior to, or at, the point of referral, e.g. if a selective serotonin reuptake inhibitor (SSRI) had been documented in a referral letter from primary care. Thus, each survival analysis describes the rate of initiation of a given treatment for patients with no available documented mention of that treatment before the relevant CPFT referral commenced.

Cox proportional hazard (CPH) models were used to examine predictors of treatment initiation. Service type at referral (relating to age), sex, ethnicity, and IMD fraction were treated as time-independent predictors. The presence of each lifetime psychiatric comorbidity was treated as a time-independent binary variable (true vs false). Confounds such as drug intolerances, allergies, other conditions prohibiting the use of certain treatments, and patient preferences were not accounted for, due to the lack of accessible data. Multicollinearity across predictor variables was assessed using the variance inflation factor (VIF). VIF ≥10 indicates a sign of severe or serious multicollinearity (25). In this study, all models had a maximum of 1.29, indicating minimal multicollinearity.

Statistical analyses were performed using R (version 4.4.0). Statistical significance was generally defined as p < α = 0.05. When summarizing models for multiple treatments, Šidák corrections were applied to α for a family of 16 models; individual uncorrected models are also presented in **Supplementary Results**. Results are reported following the Strengthening the Reporting of Observational Studies in Epidemiology checklist for cohort studies (26).

For statistical disclosure control purposes, results have been suppressed for groups (or inferrable composite groups) smaller than 10.

## 3. RESULTS

### 3.1 Demographics

The cohort (**Table 1**) included 9,083 patients, of which 59% were female. The majority of patients were categorised as White, with 16.3% having unknown ethnicity. The largest proportion were treated within working-age adult (henceforth “adult”) services (57.7%). Overall, 47.4% of the cohort had a lifetime comorbid psychiatric diagnosis, with anxiety disorders/stress reactions being the most common among CAMH (39.9%) and adult populations (33%), and organic mental disorders amongst older adults (34.7%). The timing of the recording of comorbidities, relative to that of depressive disorders, varied widely (**Supplementary Table 1**), but with a median of 0 (simultaneous coding) (see also **Supplementary Results**). Rates of any lifetime psychiatric diagnosis and MHA detentions also differed across service directorates, with the highest prevalence observed in the CAMH group (**Table 1**).

**Table 1.** Demographics, lifetime psychiatric comorbidities, and basic outcomes for patients with depressive disorders referred to CPFT. Subgroups by age at referral.

| Variable † | All | CAMH (0–16 y) | Adult (17–64 y) | Older adult (65+ y) | Comparison (A between age groups, VP versus local population) ‡ |
| --- | --- | --- | --- | --- | --- |
| Count | 9,083 (100%) | 247 (2.72%) | 5,244 (57.7%) | 3,592 (39.5%) | – |
| **Demographics** |  |  |  |  |  |
| Sex (female) | 5,356 (59%) | 200 (81%) | 2,982 (56.9%) | 2,174 (60.5%) | A: $\chi^2_2 = 62.6$, $p = 2.55 \times 10^{-14}$ ***** |
| Age (y) | 53.5 (SD 22.8) | 15.0 (SD 1.33) | 38.9 (SD 13.3) | 77.6 (SD 8.04) | – |
| Ethnicity (local population: Asian 7.88%, Black 2.07%, mixed 3.02%, other 1.66%, white 85.4%) § | Asian: 200 (2.2%)<br>Black: 67 (0.738%)<br>Mixed: 110 (1.21%)<br>Other: 100 (1.1%)<br>White: 7,125 (78.4%)<br>Unknown: 1,481 (16.3%) | Asian: < 10 (< 4.05%) †<br>Black: < 10 (< 4.05%) †<br>Mixed: 11 (4.45%)<br>Other: < 10 (< 4.05%) †<br>White: 202 (81.8%)<br>Unknown: 26 (10.5%) | Asian: 164 (3.13%)<br>Black: 53 (1.01%)<br>Mixed: 80 (1.53%)<br>Other: 84 (1.6%)<br>White: 4,122 (78.6%)<br>Unknown: 741 (14.1%) | Asian: 33 (0.919%)<br>Black: 13 (0.362%)<br>Mixed: 19 (0.529%)<br>Other: 12 (0.334%)<br>White: 2,801 (78%)<br>Unknown: 714 (19.9%) | VP: $\chi^2_4 = 447$, $p < 2.20 \times 10^{-16}$ ***** |
| IMD (Q1 most deprived, Q4 least; local population approx. Q1 13.6%, Q2 19.9%, Q3 29%, Q4 37.5%) ¶ | Q1: 1,122 (12.4%)<br>Q2: 2,090 (23%)<br>Q3: 2,759 (30.4%)<br>Q4: 2,987 (32.9%)<br>Unknown: 125 (1.38%) | Q1: 21 (8.5%)<br>Q2: 55 (22.3%)<br>Q3: 80 (32.4%)<br>Q4: 89 (36%)<br>Unknown: < 10 (< 4.05%) † | Q1: 838 (16%)<br>Q2: 1,225 (23.4%)<br>Q3: 1,555 (29.7%)<br>Q4: 1,524 (29.1%)<br>Unknown: 102 (1.95%) | Q1: 263 (7.32%)<br>Q2: 810 (22.6%)<br>Q3: 1,124 (31.3%)<br>Q4: 1,374 (38.3%)<br>Unknown: 21 (0.585%) | VP: $\chi^2_3 = 111$, $p < 2.20 \times 10^{-16}$ ***** |
| **Lifetime psychiatric comorbidities (by coded ICD-10 diagnosis)** |  |  |  |  |  |
| Organic mental disorders (F0) | 1,334 to 1,343 (14.7% to 14.8%) † | < 10 (< 4.05%) † | 88 (1.68%) | 1,246 (34.7%) | A: $\chi^2_2 = 1.90 \times 10^3$, $p < 2.20 \times 10^{-16}$ ***** |
| Substance misuse (F1) | 609 to 618 (6.7% to 6.8%) † | < 10 (< 4.05%) † | 505 (9.63%) | 104 (2.9%) | A: $\chi^2_2 = 162$, $p < 2.20 \times 10^{-16}$ ***** |
| Psychotic disorders (F2) | 294 to 303 (3.24% to 3.34%) † | < 10 (< 4.05%) † | 230 (4.39%) | 64 (1.78%) | A: $\chi^2_2 = 46.7$, $p = 7.25 \times 10^{-11}$ ***** |
| Manic episode and bipolar disorder (F30–F31) | 564 to 573 (6.21% to 6.31%) † | < 10 (< 4.05%) † | 426 (8.12%) | 138 (3.84%) | A: $\chi^2_2 = 71.5$, $p = 3.01 \times 10^{-16}$ ***** |
| Anxiety disorders and stress reactions (F40–F43) | 2,703 (29.8%) | 97 (39.3%) | 1,730 (33%) | 876 (24.4%) | A: $\chi^2_2 = 86.5$, $p < 2.20 \times 10^{-16}$ ***** |
| Eating disorders (F50) | 245 to 254 (2.7% to 2.8%) † | 53 (21.5%) | 192 (3.66%) | < 10 (< 0.278%) † | A: $\chi^2_2 = 424$, $p < 2.20 \times 10^{-16}$ ***** |
| Personality disorders (F60–F61) | 760 (8.37%) | 29 (11.7%) | 672 (12.8%) | 59 (1.64%) | A: $\chi^2_2 = 351$, $p < 2.20 \times 10^{-16}$ ***** |
| Developmental disorders (F8) | 144 (1.59%) | 12 (4.86%) | 128 (2.44%) | < 10 (< 0.278%) † | – |
| Intellectual disability (F7) | 33 to 51 (0.363% to 0.561%) † | < 10 (< 4.05%) † | 33 (0.629%) | < 10 (< 0.278%) † | – |
| Childhood behavioural and emotional disorders (F9) | 156 (1.72%) | 31 (12.6%) | 109 (2.08%) | 16 (0.445%) | A: $\chi^2_2 = 210$, $p < 2.20 \times 10^{-16}$ ***** |
| Any psychiatric comorbidity (F, excluding depression as F20.4, F25.1, F31.3–F31.5, F32–F33, F38.1, F41.2) | 4,301 (47.4%) | 148 (59.9%) | 2,508 (47.8%) | 1,645 (45.8%) | A: $\chi^2_2 = 19.6$, $p = 5.52 \times 10^{-5}$ **** |
| Intentional self-harm (X60–X84) | 405 to 414 (4.46% to 4.56%) † | 12 (4.86%) | 282 (5.38%) | 123 (3.42%) | A: $\chi^2_2 = 18.6$, $p = 9.10 \times 10^{-5}$ **** |
| **Episodes and outcomes** |  |  |  |  |  |
| Number with index care episodes spanning diagnosis, closely following diagnosis, and closely preceding (ending with) diagnosis, respectively | 8,895 (97.9%), 91 (1%), 97 (1.07%) | 243 (98.4%), < 10 (< 4.05%) †, < 10 (< 4.05%) † | 5,135 (97.9%), 40 (0.763%), 69 (1.32%) | 3,517 (97.9%), 51 (1.42%), 24 (0.668%) | – |
| Duration with services for index episode of care (days) | median 192 (range 1 to 3,020) | median 680 (range 8 to 2,237) | median 219 (range 1 to 3,020) | median 146 (range 1 to 2,675) | See other table |
| Ever detained under the Mental Health Act at/after start of index episode | 482 (5.31%) | 22 (8.91%) | 319 (6.08%) | 141 (3.93%) | A: $\chi^2_2 = 26.3$, $p = 1.94 \times 10^{-6}$ ***** |
| Number with any HoNOS measurement for index episode | 6,136 (67.6%) | 44 (17.8%) | 3,841 (73.2%) | 2,251 (62.7%) | A: $\chi^2_2 = 395$, $p < 2.20 \times 10^{-16}$ ***** |
| Number with two HoNOS scores at least 7 days apart | 3,317 (36.5%) | 26 (10.5%) | 1,888 (36%) | 1,403 (39.1%) | A: $\chi^2_2 = 82.6$, $p < 2.20 \times 10^{-16}$ ***** |
| Time between HoNOS scores (days) ¶ | median 279 (range 7 to 2,652) | median 364 (range 173 to 1,426) | median 307 (range 7 to 2,652) | median 245 (range 7 to 2,596) | – |
| HoNOS total score: first (0–48) | 11.9 (SD 5.10) | 13.0 (SD 5.60) | 12.6 (SD 5.21) | 10.6 (SD 4.62) | A: $F_{2,6133} = 118$, $p < 2.20 \times 10^{-16}$ ***** |
| HoNOS depression score: first (0–4) | 2.32 (SD 0.906) | 2.11 (SD 0.754) | 2.43 (SD 0.870) | 2.12 (SD 0.934) | A: $F_{2,6133} = 87.1$, $p < 2.20 \times 10^{-16}$ ***** |
| HoNOS total score: change (last – first) ¶ | –2.99 (SD 5.63) | –3.27 (SD 5.01) | –3.23 (SD 6.09) | –2.66 (SD 4.93) | See other table |
| HoNOS depression score: change (last – first) ¶ | –0.850 (SD 1.18) | –0.385 (SD 0.983) | –0.777 (SD 1.21) | –0.956 (SD 1.13) | See other table |
| Died during follow-up | 313 to 322 (3.45% to 3.55%) † | < 10 (< 4.05%) † | 64 (1.22%) | 249 (6.93%) | A: $\chi^2_2 = 213$, $p < 2.20 \times 10^{-16}$ ***** |

†Small number suppression applied. Percentages are of the relevant age group, except for overall counts, which are of the overall total. SD, standard deviation. ‡Comparisons to population frequencies use all subjects excluding unknown values, and unknown values may be biased; \*\*\*\*\* $p < 0.00001$ ; \*\*\*\* $p < 0.0001$ ; \*\*\* $p < 0.001$ ; \*\* $p < 0.01$ ; \* $p < 0.05$ ; NS, not significant. §Cambridgeshire & Peterborough (C&P), https://cambridgeshireinsight.org.uk/, Census 2021 data, retrieved 2024-07-04. ¶Based on C&P Lower layer Super Output Areas (LSOAs), assumed to have approximately equal populations. ¶For those with two HoNOS measurements at least 7 days apart.

### 3.2 Predictors of initial symptom severity and duration with services

The median length of time spent with services was 192 days, varying across directorates (**Table 1**). On average, patients spent significantly longer with services if they were in the CAMH age band, female, or more socio-economically deprived, and less time if they were aged 65+ (**Supplementary Table 2**). Most psychiatric comorbidities were associated with longer time with services, except for substance misuse, and self-harm was also associated with shorter duration (**Supplementary Table 2**).

Based on initial HoNOS scores, depressive symptoms were more severe among adult patients, relative to other age groups (**Table 1; Supplementary Table 3**). Depressive symptom scores were unaffected by sex, or ethnicity (except as noted below), and where there were associations with psychiatric comorbidities, depressive symptoms were less severe when a comorbidity was present, perhaps reflecting situations where depression was not the dominant problem (**Supplementary Table 3**). Greater severity of total HoNOS scores was associated with male sex, greater socio-economic deprivation, and some comorbidities (substance misuse, eating disorders, personality disorders, and developmental disorders), although BPAD was associated with lesser total severity scores (**Supplementary Table 3**).

Those with unknown ethnicity spent less time with services (**Supplementary Table 2**) and experienced lesser severity of depressive symptoms (**Supplementary Table 3**), likely reflecting reverse causality (under-coding of ethnicity when patients spent short periods with services) (see also **Supplementary Results**).

### 3.4 Treatment initiation

Figure 2 illustrates the variation in initiation of different treatments. Nearly all patients (97%) were prescribed a new antidepressant medication (new as far as could be ascertained from the data), including augmenting agents such as SGAs, within one year of referral. SSRIs were most commonly prescribed (∼74%), followed by mirtazapine and antipsychotics (compatible with NICE augmentation guidance). Initiation of these treatments peaked within the first year, then plateaued or increased slightly thereafter. Tricyclic antidepressants (TCAs), lithium, and lamotrigine were less frequently prescribed: after four years, about a quarter or less had had each of these treatments initiated. MAOIs and ECT were rarely initiated, and likewise trazodone, flupentixol, or triiodothyronine. Approximately half of patients received psychological therapy (in secondary care) within the first year. For crisis team use and admissions, see also Supplementary Results.

**Figure 2.**
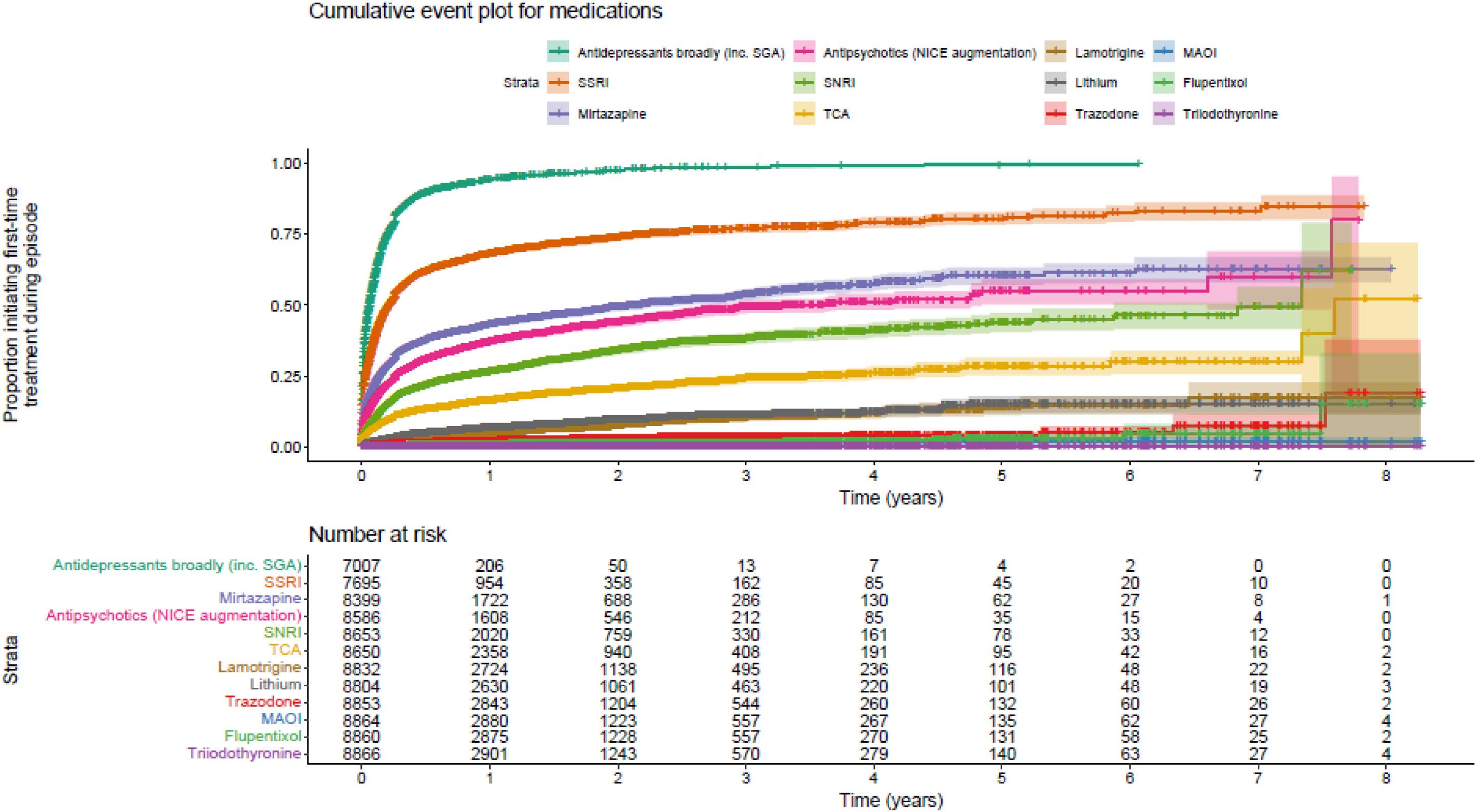
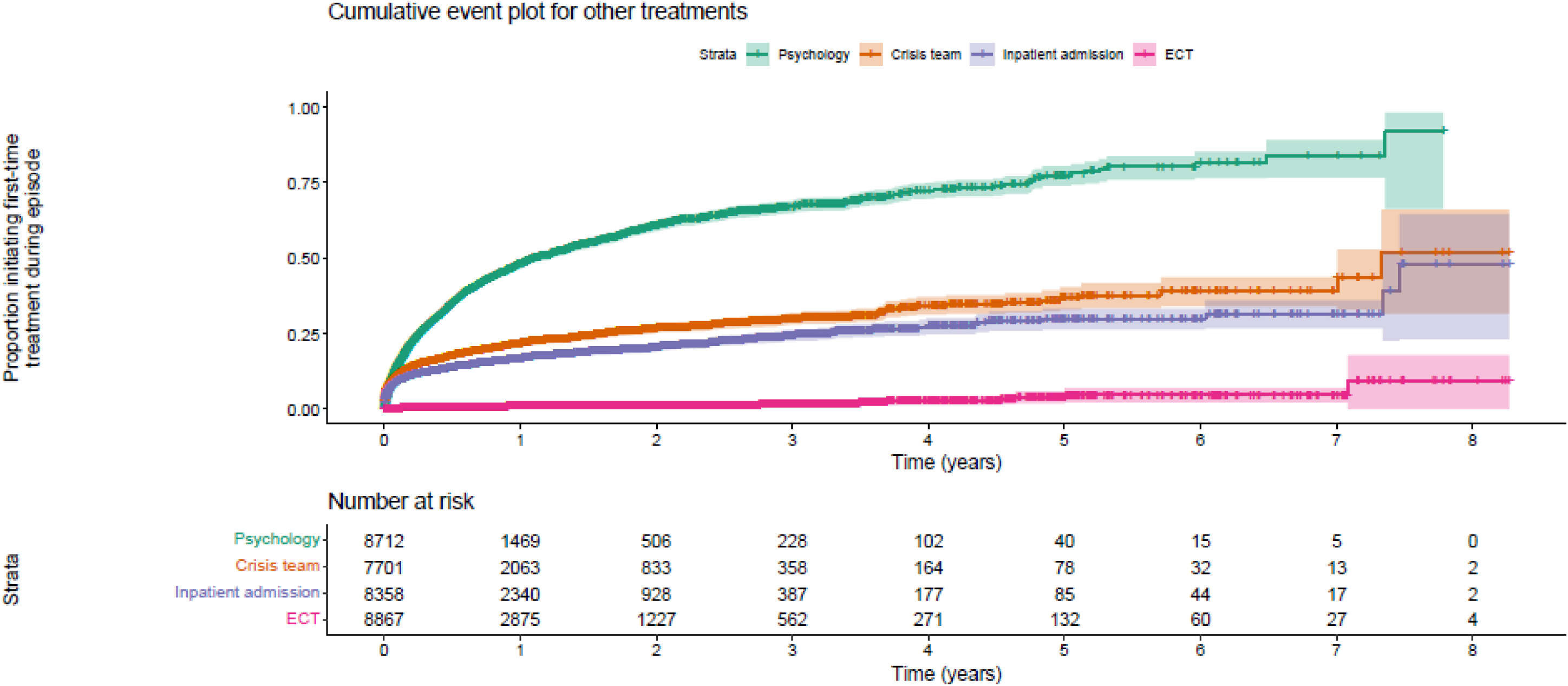
Cumulative event plots for medications and other treatments. Participants were only analysed when they had no prior detectable reference to the treatment in question before/at time 0 (the start of the index episode); this accounts for the variation in the number at risk at this time. **Upper panels:** medications. **Lower panels:** other treatments. (ECT, electroconvulsive therapy; MAOI; monoamine oxidase inhibitor; NICE, UK National Institute for Health and Care Excellence; SGA, second-generation antipsychotic; SNRI, serotonin/noradrenaline reuptake inhibitor; SSRI, selective serotonin reuptake inbitor; TCA, tricyclic antidepressant.)

### 3.5 Predictors of treatment

Compared to patients of working-age adult services, children and older adults were less likely to be prescribed any type of antidepressant collectively, but were more likely to receive psychological therapy in secondary care (**Supplementary Table 4,** summary corrected for multiple comparisons; **Supplementary Tables 5–20**, underlying analyses). Specific drug patterns differed by age group and comorbidities, but aligned broadly with clinical guidelines (**Supplementary Table 4**).

Males had higher rates of inpatient admissions and initiating mirtazapine, whereas females were more commonly prescribed TCAs. Patients living in more deprived areas were more likely to be admitted to hospital, but less likely to be prescribed lithium. The CAMH group were also more likely to be admitted to hospital, while older adults were less so. There were no ethnicity effects predicting treatment except those relating to unknown ethnicity, discussed above (**Supplementary Table 4**), and the isolated result (surviving correction for multiple comparisons) that patients of mixed ethnicity were more likely than others to be prescribed trazodone.

The **Supplementary Results** expand on specific treatment types and comorbidities.

### 3.6 Predictors of change in symptoms

Across all patients, depression and total HoNOS scores improved significantly across the episode of care (**Table 1**; **Table 2**). Lesser improvements, in both depression and total scores, were seen in patients with greater socio-economic deprivation, or psychiatric comorbidities including organic mental disorders, substance misuse, and personality disorders. Lesser improvement in depression sub-scores was associated with comorbid BPAD, or anxiety disorders/stress reactions. Depression sub-scores improved significantly more for older adults compared to working-age adults. Total scores improved more for those with comorbid psychotic disorders (**Table 2**; see also **Supplementary Results**).

**Table 2.** Predictors of change in HoNOS scores (last minus first, more negative changes are better), for those with two such scores at least 7 days apart.

| Term | *F* (via Type III sums of squares) | *p<sub>F</sub>* | Level | Coefficient (95% CI) | Standard error | *t* | *p<sub>t</sub>* |
| --- | --- | --- | --- | --- | --- | --- | --- |
| **DEPRESSION SUB-SCORES** |  |  |  |  |  |  |  |
| (Intercept) |  |  | – | –0.654 (CI –0.801 to –0.507) | 0.0751 | *t*<sub>3246</sub> = –8.71 | *p* < 2.20 × 10<sup>–16</sup> ***** |
| Service (age band) at referral | *F*<sub>2,3246</sub> = 8.16 | *p* = 0.000293 *** | Adult | Reference |  |  |  |
|  |  |  | CAMHS | +0.344 (CI –0.118 to +0.806) | 0.236 | *t*<sub>3246</sub> = +1.46 | *p* = 0.144, NS |
|  |  |  | Older adult | –0.189 (CI –0.289 to –0.0881) | 0.0513 | *t*<sub>3246</sub> = –3.68 | *p* = 0.000237 *** |
| Sex | *F* < 1 | NS |  |  |  |  |  |
| Ethnicity | *F*<sub>5,3246</sub> = 1.67 | *p* = 0.138, NS |  |  |  |  |  |
| IMD fraction (0–1, higher values indicating less deprived) | *F*<sub>1,3246</sub> = 7.50 | *p* = 0.00620 ** | – | –0.214 (CI –0.368 to –0.0609) | 0.0782 | *t*<sub>3246</sub> = –2.74 | *p* = 0.00620 ** |
| Organic mental disorders (F0) | *F*<sub>1,3246</sub> = 27.6 | *p* = 1.57 × 10<sup>–7</sup> ***** | True | +0.332 (CI +0.208 to +0.456) | 0.0633 | *t*<sub>3246</sub> = +5.26 | *p* = 1.57 × 10<sup>–7</sup> ***** |
| Substance misuse (F1) | *F*<sub>1,3246</sub> = 10.4 | *p* = 0.00126 ** | True | +0.272 (CI +0.107 to +0.438) | 0.0844 | *t*<sub>3246</sub> = +3.23 | *p* = 0.00126 ** |
| Psychotic disorders (F2) | *F* < 1 | NS |  |  |  |  |  |
| Manic episode and bipolar disorder (F30–F31) | *F*<sub>1,3246</sub> = 6.09 | *p* = 0.0137 * | True | +0.193 (CI +0.0397 to +0.347) | 0.0783 | *t*<sub>3246</sub> = +2.47 | *p* = 0.0137 * |
| Anxiety disorders and stress reactions (F40–F43) | *F*<sub>1,3246</sub> = 5.50 | *p* = 0.0190 * | True | +0.103 (CI +0.0169 to +0.189) | 0.0439 | *t*<sub>3246</sub> = +2.35 | *p* = 0.0190 * |
| Eating disorders (F50) | *F*<sub>1,3246</sub> = 2.77 | *p* = 0.0964, NS |  |  |  |  |  |
| Personality disorders (F60–F61) | *F*<sub>1,3246</sub> = 34.2 | *p* = 5.42 × 10<sup>–9</sup> ***** | True | +0.403 (CI +0.268 to +0.538) | 0.0689 | *t*<sub>3246</sub> = +5.85 | *p* = 5.42 × 10<sup>–9</sup> ***** |
| Intellectual disability (F7) | *F*<sub>1,3246</sub> = 1.21 | *p* = 0.272, NS |  |  |  |  |  |
| Developmental disorders (F8) | *F* < 1 | NS |  |  |  |  |  |
| Childhood behavioural and emotional disorders (F9) | *F*<sub>1,3246</sub> = 2.09 | *p* = 0.148, NS |  |  |  |  |  |
| Intentional self-harm (X60–X84) | *F*<sub>1,3246</sub> = 2.26 | *p* = 0.133, NS |  |  |  |  |  |
| Selective serotonin reuptake inhibitor (SSRI) | *F*<sub>1,3246</sub> = 10.8 | *p* = 0.00104 ** | True | –0.144 (CI –0.230 to –0.0581) | 0.0439 | *t*<sub>3246</sub> = –3.28 | *p* = 0.00104 ** |
| Tricyclic antidepressant (TCA) | *F* < 1 | NS |  |  |  |  |  |
| Monoamine oxidase inhibitor (MAOI) | *F*<sub>1,3246</sub> = 1.49 | *p* = 0.223, NS |  |  |  |  |  |
| Serotonin/noradrenaline reuptake inhibitor (SNRI) | *F* < 1 | NS |  |  |  |  |  |
| Mirtazapine | *F* < 1 | NS |  |  |  |  |  |
| Trazodone | *F* < 1 | NS |  |  |  |  |  |
| Antipsychotics endorsed by NICE for depression | *F* < 1 | NS |  |  |  |  |  |

| Term | $F$ (via Type III sums of squares) | $p_F$ | Level | Coefficient (95% CI) | Standard error | $t$ | $p_t$ |
| --- | --- | --- | --- | --- | --- | --- | --- |
| Flupentixol | $F < 1$ | NS | | | | | |
| Lithium | $F_{1,3246} = 5.25$ | $p = 0.0220^*$ | True | -0.179 (CI -0.333 to -0.0258) | 0.0783 | $t_{3246} = -2.29$ | $p = 0.0220^*$ |
| Lamotrigine | $F < 1$ | NS | | | | | |
| Electroconvulsive therapy (ECT) | $F_{1,3246} = 1.41$ | $p = 0.235$, NS | | | | | |
| Psychology input | $F_{1,3246} = 2.24$ | $p = 0.134$, NS | | | | | |
| **TOTAL SCORES** |  |  |  |  |  |  |  |
| (Intercept) | | | – | -3.01 (CI -3.71 to -2.32) | 0.355 | $t_{3246} = -8.50$ | $p < 2.20 \times 10^{-16}$ ***** |
| Service (age band) at referral | $F_{2,3246} = 1.30$ | $p = 0.274$, NS | | | | | |
| Sex | $F < 1$ | NS | | | | | |
| Ethnicity | $F_{5,3246} = 1.46$ | $p = 0.201$, NS | | | | | |
| IMD fraction (0–1, higher values indicating less deprived) | $F_{1,3246} = 4.32$ | $p = 0.0378^*$ | – | -0.768 (CI -1.49 to -0.0431) | 0.370 | $t_{3246} = -2.08$ | $p = 0.0378^*$ |
| Organic mental disorders (F0) | $F_{1,3246} = 37.2$ | $p = 1.19 \times 10^{-9}$ ***** | True | +1.82 (CI +1.24 to +2.41) | 0.299 | $t_{3246} = +6.10$ | $p = 1.19 \times 10^{-9}$ ***** |
| Substance misuse (F1) | $F_{1,3246} = 8.89$ | $p = 0.00289$ ** | True | +1.19 (CI +0.407 to +1.97) | 0.399 | $t_{3246} = +2.98$ | $p = 0.00289$ ** |
| Psychotic disorders (F2) | $F_{1,3246} = 7.27$ | $p = 0.00705$ ** | True | -1.24 (CI -2.14 to -0.338) | 0.460 | $t_{3246} = -2.70$ | $p = 0.00705$ ** |
| Manic episode and bipolar disorder (F30–F31) | $F_{1,3246} = 1.61$ | $p = 0.204$, NS | | | | | |
| Anxiety disorders and stress reactions (F40–F43) | $F < 1$ | NS | | | | | |
| Eating disorders (F50) | $F < 1$ | NS | | | | | |
| Personality disorders (F60–F61) | $F_{1,3246} = 68.8$ | $p < 2.20 \times 10^{-16}$ ***** | True | +2.70 (CI +2.06 to +3.34) | 0.326 | $t_{3246} = +8.29$ | $p < 2.20 \times 10^{-16}$ ***** |
| Intellectual disability (F7) | $F_{1,3246} = 6.46$ | $p = 0.0111^*$ | True | +8.07 (CI +1.84 to +14.3) | 3.18 | $t_{3246} = +2.54$ | $p = 0.0111^*$ |
| Developmental disorders (F8) | $F < 1$ | NS | | | | | |
| Childhood behavioural and emotional disorders (F9) | $F < 1$ | NS | | | | | |
| Intentional self-harm (X60–X84) | $F_{1,3246} = 1.10$ | $p = 0.295$, NS | | | | | |
| Selective serotonin reuptake inhibitor (SSRI) | $F_{1,3246} = 1.27$ | $p = 0.260$, NS | | | | | |
| Tricyclic antidepressant (TCA) | $F_{1,3246} = 5.58$ | $p = 0.0183^*$ | True | +0.610 (CI +0.103 to +1.12) | 0.258 | $t_{3246} = +2.36$ | $p = 0.0183^*$ |
| Monoamine oxidase inhibitor (MAOI) | $F_{1,3246} = 4.45$ | $p = 0.0349^*$ | True | +2.17 (CI +0.153 to +4.18) | 1.03 | $t_{3246} = +2.11$ | $p = 0.0349^*$ |
| Serotonin/noradrenaline reuptake inhibitor (SNRI) | $F < 1$ | NS | | | | | |
| Mirtazapine | $F < 1$ | NS | | | | | |
| Trazodone | $F < 1$ | NS | | | | | |
| Antipsychotics endorsed by NICE for depression | $F_{1,3246} = 7.42$ | $p = 0.00650$<br>** | True | -0.606 (CI -1.04 to -0.170) | 0.223 | $t_{3246} = -2.72$ | $p = 0.00650$<br>** |
| Flupentixol | $F < 1$ | NS | | | | | |
| Lithium | $F < 1$ | NS | | | | | |
| Lamotrigine | $F_{1,3246} = 5.41$ | $p = 0.0201$ * | True | +0.996 (CI +0.156 to +1.84) | 0.428 | $t_{3246} = +2.33$ | $p = 0.0201$ * |
| Electroconvulsive therapy (ECT) | $F_{1,3246} = 1.05$ | $p = 0.305$,<br>NS | | | | | |
| Psychology input | $F < 1$ | NS | | | | | |

In relation to treatments, depression sub-scores reduced significantly more than the average for patients receiving SSRIs or lithium (**Table 2**). Use of antipsychotics endorsed by NICE for depression was associated with significantly improved overall HoNOS scores (over and above the presence of coded psychotic disorders from the F2 group, and other predictors), while treatment with a TCA, MAOI, or lamotrigine was associated with lesser improvement in overall scores (**Table 2**). There is of course potential for confounding by indication. All these effects remained following the sensitivity analysis excluding patients with BPAD, except the associations between IMD or MAOIs and overall HoNOS scores, and between lithium and depression sub-scores (**Supplementary Table 21**).

## 4. DISCUSSION

We characterised a cohort of patients with depressive disorders receiving treatment from secondary MH care services, along with a longitudinal analysis of the treatments they received. Almost half of the cohort suffered with at least one psychiatric comorbidity, reflecting the complex challenges patients face. Treatment patterns differed across age groups, sex/gender, socio-economic status, and psychiatric comorbidities. Differences relating to age group and comorbidity aligned broadly with established clinical guidelines; however, potential treatment gaps remained. Despite national guidance, the application of some further-line treatments was limited, and treatment outcomes were worse for those with comorbidities or lower socio-economic status.

### 4.1 Patient characteristics

Our cohort had a higher proportion of females than males, consistent with previous studies (27,28). We expand on existing data and confirm the high prevalence of psychiatric comorbidities among patients with persistent depression (7) and greater treatment resistance (3,6). These findings illustrate the multifaceted challenges patients face, supporting what some authors have interpreted as a general psychopathology dimension, the p-factor (29), advocating for a more transdiagnostic approach to treatment.

Comorbidities were less common than in similar studies (5,6,12,16), perhaps due to under-coding, despite previous research excluding patients experiencing specific comorbidities (e.g. schizophrenia, bipolar disorder, dementia, eating disorders, substance misuse). Furthermore, including patients with organic mental disorders, encompassing dementia, may have raised the average age of our cohort compared to other studies. We provide a comprehensive account reflecting the complexity of cases treated in secondary care.

While depressive symptom scores were most severe in working-age adults, comorbidities and rates of psychiatric admission and MHA detention were highest among the CAMH group. There is some potential for unmeasured coding bias, e.g. if the likelihood of diagnostic coding were less for adults not requiring admission, or for children not requiring medication. Three-quarters of adult mental illness manifests by age 24 (30); with an increasing demand for CAMH services, there is an urgency for early intervention among young people experiencing depression. Children and young people with complex needs, such as those in our sample, are at marked risk of persistent problems (31).

### 4.2 Treatment initiation and outcomes

Established treatments for persistent depression, including SSRIs and SGAs, were routinely prescribed, and these appeared particularly effective for treating depressive and/or overall symptoms. There is potential for confounding by indication, e.g. if these drug classes were prescribed disproportionately for “less refractory” or earlier-stage depression; however, while this is plausible for SSRIs (a first-line treatment), it is less plausible as an interpretation for SGAs (usually a later-stage augmentation agent, or one prescribed for an alternative comorbidity). However, other further-line treatments including lithium, lamotrigine, TCAs, MAOIs, and ECT, were used infrequently, despite their proven efficacy. Similar treatment patterns, including under-use of further-line treatments, have been reported nationally (17) and internationally (32). Integrating the views of clinicians, patients, and commissioning groups influencing prescribing could help explain these treatment gaps and direct future service improvements. Notably, the apparent lesser impact of TCAs and MAOIs may have been confounded by the severity of depression in patients receiving these treatments.

Low use of lithium may be partially explained by the blood tests it requires (33). Lithium use was inversely associated with socio-economic status indicators (and lower socio-economic status was also associated with greater severity of depression). It is unlikely that greater severity drove lesser lithium use, but is unclear if a relative reluctance to use lithium in this group was driven by patients (e.g. relative deprivation making phlebotomy attendance harder), by clinicians (e.g. perceiving such patients as less able to cope with the requirements of lithium), or both.

Combining psychotherapy and pharmacotherapy therapy is superior to either modality alone for persistent depression (34), yet approximately half of our cohort received psychological input (detectable within secondary care) within the first year of being treated in secondary care. Any psychological input in primary care was unmeasured. In comparison, a study recruiting TRD participants from primary and secondary care found that half reported completing an adequate course of psychological therapy (12), suggesting a treatment gap. Notably, current definitions of TRD neglect the role of psychotherapy; it may therefore seem unmerited to label patients as “treatment-resistant” without considering this option (35).

The presence of comorbidities and lower socio-economic status indicators were associated with worse treatment outcomes, echoing previous research (3). We examined each comorbidity individually, confirming comorbid anxiety disorders/stress reactions (36) and personality disorders (7) to be strong predictors of poor treatment response. Patients with comorbid substance misuse experienced especially poor outcomes; these patients spent significantly less time with services, despite national guidance on treating the two entities together (37).

The relationship between socio-economic status and persistent depression is corroborated by previous studies in primary care (38) and trial settings (39). Further work is needed to understand the mechanisms preventing these patients from benefiting from current practice. The composite IMD measure encompasses income, employment, education, crime, barriers to housing, and living environment. It may be a proxy for adversities impacting not only overall illness severity, but also the effectiveness of current treatments. Future work is needed to understand the barriers to improvement these patients face.

### 4.4 Improving future outcomes

The treatment gaps suggested by this study signal a need for more effective strategies across secondary care services. Evidence on how to implement clinical guidelines more effectively is scarce, but a multicentre randomised controlled trial of a specialist depression service (SDS) in secondary care, offering NICE-recommended collaborative psychological and pharmacological treatments, found the SDS produced significantly better long-term clinical outcomes compared to treatment as usual, underscoring the efficacy of current treatments when endorsed by suitable structure and resource. New treatments, such as repetitive transcranial magnetic stimulation (rTMS) (40), ketamine (41), and psilocybin (42) have demonstrated promising results; however, weakness of the evidence base about their cost-effectiveness impedes more widespread use in the UK (43). Thus, effective treatments are known, with more on the horizon, but implementation barriers need to be addressed to support real-world practice.

### 4.5 Strengths and limitations

Our longitudinal dataset enabled us to evaluate real-world treatment patterns over an approximately eight-year period, yielding statistical power; however, observation periods for each patient were not equal, and across the course of the dataset, changes to national guidance and service provision/resources may have influenced practice. Depression is typically more severe in a secondary care sample, so our results do not represent everyone with depression, but are likely representative of those with persistent depression using such services. We tried to identify the episode of care most closely related to the depression diagnosis (Figure 1), but in some cases where patients had a psychiatric comorbidity, depression may not have been the priority, thus the treatment focus may have been elsewhere.

The use of an observational design inevitably brings the possibility of unmeasured confounds, and confounding by indication (e.g. if treatments are decided on partly by severity, their apparent effectiveness may be confounded). We controlled observationally for a number of confounds, but these possibilities remain as a limitation.

Focusing on secondary care allowed an in-depth analysis of further-line treatments; however, without primary care data (including NHS Talking Therapies data) we were unable to interrogate the whole treatment pathway, including failed treatments and duration of symptoms. Linking patient data across primary/secondary care would generate a fuller picture. Future studies reporting encounters with secondary care for other reasons may also help to establish predictors and earlier intervention points.

The main outcome source (HoNOS score) was not available for everyone, and HoNOS recording may have been less likely in patients who were with services briefly, potentially resulting in our data reflecting more severe illness. Shorter time with services was associated with under-coding of ethnicity, reflecting a similar problem. While our dataset derived from the Cambridgeshire and Peterborough region, an economically diverse area in a mixed rural/urban setting, caution must be applied when generalising our findings.

### 4.6 Conclusions

Depression in secondary care is complex, not least because of the psychiatric comorbidities many patients face. Secondary care services are effective and improve symptoms, but there is room for improvement. Our results show not all recommended treatments are widely used, and those with lower socio-economic status indicators are at a disadvantage, experiencing greater overall illness severity and less improvement in overall or depressive symptoms. Future work is needed to target patients experiencing comorbidities and those who are socio-economically deprived.

## Data availability

The analytical code for the study is available from the corresponding author on reasonable request. Raw de-identified data are not publicly available, under the terms of NHS Research Ethics approvals; for details of access and conditions, contact.

## Acknowledgements

We thank the i-VALiD patient and public involvement group for their support and contribution. We also wish to thank Dr Catherine Hatfield for her expert clinical advice and input towards interpreting aspects of the data.

## Author contributions

CW designed the research programme this study belongs to. LH and RNC designed the study, extracted and analysed the data, and drafted the manuscript. LA, EO, and JL contributed to the development of the analytical code. All authors contributed to data interpretation, and edited and approved the final manuscript.

## References

1. Institute of Health Metrics and Evaluation. Global Health Data Exchange (GHDx) [Internet]. 2023. Available from: https://vizhub.healthdata.org/gbd-results/

2. UK House of Commons. Mental health statistics: prevalence, services and funding in England [Internet]. 2024. Available from: https://researchbriefings.files.parliament.uk/documents/SN06988/SN06988.pdf

3. Rush AJ, Trivedi MH, Wisniewski SR, Nierenberg AA, Stewart JW, Warden D, et al. Acute and Longer-Term Outcomes in Depressed Outpatients Requiring One or Several Treatment Steps: A STAR*D Report. Am J Psychiatry [Internet]. 2006 Nov;163(11):1905–17. Available from: http://psychiatryonline.org/doi/abs/10.1176/ajp.2006.163.11.1905

4. Gaynes BN, Lux L, Gartlehner G, Asher G, Forman-Hoffman V, Green J, et al. Defining treatment-resistant depression. Depress Anxiety [Internet]. 2020 Feb;37(2):134–45. Available from: http://www.ncbi.nlm.nih.gov/pubmed/31638723

5. Adekkanattu P, Olfson M, Susser LC, Patra B, Vekaria V, Coombes BJ, et al. Comorbidity and healthcare utilization in patients with treatment resistant depression: A large-scale retrospective cohort analysis using electronic health records. J Affect Disord [Internet]. 2023;324(December 2022):102–13. Available from:10.1016/j.jad.2022.12.044

6. Pappa S, Shah M, Young S, Anwar T, Ming T. Care pathways, prescribing practices and treatment outcomes in major depressive disorder and treatment-resistant depression: retrospective, population-based cohort study. BJPsych Open [Internet]. 2024 Jan 19;10(1):e32. Available from:https://www.cambridge.org/core/product/identifier/S2056472423006270/type/journal_article

7. Rost F, Booker T, Gonsard A, de Felice G, Asseburg L, Malda-Castillo J, et al. The complexity of treatment-resistant depression: A data-driven approach. J Affect Disord [Internet]. 2024 Apr; Available from:https://linkinghub.elsevier.com/retrieve/pii/S0165032724007043

8. Mrazek DA, Hornberger JC, Altar CA, Degtiar I. A Review of the Clinical, Economic, and Societal Burden of Treatment-Resistant Depression: 1996–2013. Psychiatr Serv [Internet]. 2014 Aug;65(8):977–87. Available from:https://psychiatryonline.org/doi/10.1176/appi.ps.201300059

9. National Institute for Health and Clinical Excellence. Depression in adults: treatment and management NICE guideline. NICE Guidel [Internet]. 2022;(June). Available from: www.nice.org.uk/guidance/ng222

10. Cleare A, Pariante C, Young A, Anderson I, Christmas D, Cowen P, et al. Evidence-based guidelines for treating depressive disorders with antidepressants: A revision of the 2008 British Association for Psychopharmacology guidelines. J Psychopharmacol [Internet]. 2015 May 12;29(5):459–525. Available from:https://journals.sagepub.com/doi/10.1177/0269881115581093

11. Wiles N, Taylor A, Turner N, Barnes M, Campbell J, Lewis G, et al. Management of treatment-resistant depression in primary care: a mixed-methods study. Br J Gen Pract [Internet]. 2018 Oct;68(675):e673–81. Available from:https://bjgp.org/lookup/doi/10.3399/bjgp18X699053

12. Day E, Shah R, Taylor RW, Marwood L, Nortey K, Harvey J, et al. A retrospective examination of care pathways in individuals with treatment-resistant depression. BJPsych Open [Internet]. 2021 May 14;7(3):e101. Available from:https://www.cambridge.org/core/product/identifier/S2056472421000594/type/journal_article

13. Bogowicz P, Curtis HJ, Walker AJ, Cowen P, Geddes J, Goldacre B. Trends and variation in antidepressant prescribing in English primary care: a retrospective longitudinal study. BJGP Open [Internet]. 2021 Aug;5(4):BJGPO.2021.0020. Available from: http://bjgpopen.org/lookup/doi/10.3399/BJGPO.2021.0020

14. Gillman K. “Much ado about nothing”: monoamine oxidase inhibitors, drug interactions, and dietary tyramine. CNS Spectr [Internet]. 2017 Oct 2;22(5):385–7. Available from: https://www.cambridge.org/core/product/identifier/S1092852916000651/type/journal_article

15. Heerlein K, Perugi G, Otte C, Frodl T, Degraeve G, Hagedoorn W, et al. Real-world evidence from a European cohort study of patients with treatment resistant depression: Treatment patterns and clinical outcomes. J Affect Disord [Internet]. 2021 Jul;290:334–44. Available from: https://linkinghub.elsevier.com/retrieve/pii/S0165032721003086

16. Lundberg J, Cars T, Lööv S-Å, Söderling J, Sundström J, Tiihonen J, et al. Association of Treatment-Resistant Depression With Patient Outcomes and Health Care Resource Utilization in a Population-Wide Study. JAMA Psychiatry [Internet]. 2023 Feb 1;80(2):167. Available from: https://jamanetwork.com/journals/jamapsychiatry/fullarticle/2799488

17. Costa T, Menzat B, Engelthaler T, Fell B, Franarin T, Roque G, et al. The burden associated with, and management of, difficult-to-treat depression in patients under specialist psychiatric care in the United Kingdom. J Psychopharmacol [Internet]. 2022 May 4;36(5):545–56. Available from:https://journals.sagepub.com/doi/10.1177/02698811221090628

18. Chapman N, Browning M, Baghurst D, Hotopf M, Willis D, Haylock S, et al. Setting national research priorities for difficult-to-treat depression in the UK between 2021-2026. J Glob Health [Internet]. 2022 Dec 22;12:09004. Available from:https://jogh.org/2022/jogh-12-09004

19. World Health Organization. The ICD-10 classification of mental and behavioural disorders: clinical descriptions and diagnostic guidelines (Vol. 1). 1992.

20. UK Public General Acts. Mental Heath Act 1983 [Internet]. UK Government; 1983. Available from: https://www.legislation.gov.uk/ukpga/1983/20

21. Sultana J, Chang CK, Hayes RD, Broadbent M, Stewart R, Corbett A, et al. Associations between risk of mortality and atypical antipsychotic use in vascular dementia: a clinical cohort study. Int J Geriatr Psychiatry [Internet]. 2014 Dec 14;29(12):1249–54. Available from: https://onlinelibrary.wiley.com/doi/10.1002/gps.4101

22. Cleare A, Pariante C, Young A, Anderson I, Christmas D, Cowen P, et al. Evidence-based guidelines for treating depressive disorders with antidepressants: A revision of the 2008 British Association for Psychopharmacology guidelines. J Psychopharmacol [Internet]. 2015 May 12;29(5):459–525. Available from:http://journals.sagepub.com/doi/10.1177/0269881115581093

23. Wing JK, Curtis RH, Beevor AS. Health of the Nation Outcome Scales (HoNOS). Br J Psychiatry [Internet]. 1999 May 3;174(5):432–4. Available from:https://www.cambridge.org/core/product/identifier/S0007125000262302/type/journal_article

24. Cambridgeshire & Peterborough Insight. Cambridgeshire & Peterborough Insight [Internet]. 2024. Available from:https://cambridgeshireinsight.org.uk/population/#/view-report/8a66021c1b2b41fb984533efc510ab4a/aFirstFeature/G28

25. O’brien RM. A Caution Regarding Rules of Thumb for Variance Inflation Factors. Qual Quant [Internet]. 2007 Sep 11;41(5):673–90. Available from:http://link.springer.com/10.1007/s11135-006-9018-6

26. STROBE. STROBE Checklist for cohort studies [Internet]. 2007. Available from: https://www.strobe-statement.org/fileadmin/Strobe/uploads/checklists/STROBE-checklist-v4-cohort.pdf

27. Heerlein K, Young AH, Otte C, Frodl T, Degraeve G, Hagedoorn W, et al. Real-world evidence from a European cohort study of patients with treatment resistant depression: Baseline patient characteristics. J Affect Disord [Internet]. 2021 Mar;283:115–22. Available from: https://linkinghub.elsevier.com/retrieve/pii/S0165032720330585

28. Hollon SD, Shelton RC, Wisniewski S, Warden D, Biggs MM, Friedman ES, et al. Presenting characteristics of depressed outpatients as a function of recurrence: Preliminary findings from the STAR*D clinical trial. J Psychiatr Res [Internet]. 2006 Feb;40(1):59–69. Available from: https://linkinghub.elsevier.com/retrieve/pii/S0022395605000920

29. Caspi A, Houts RM, Belsky DW, Goldman-Mellor SJ, Harrington H, Israel S, et al. The p Factor. Clin Psychol Sci [Internet]. 2014 Mar 14;2(2):119–37. Available from: https://journals.sagepub.com/doi/10.1177/2167702613497473

30. Kessler RC, Berglund P, Demler O, Jin R, Merikangas KR, Walters EE. Lifetime prevalence and age-of-onset distributions of DSM-IV disorders in the National Comorbidity Survey Replication. Arch Gen Psychiatry [Internet]. 2005 Jun;62(6):593–602. Available from: http://www.ncbi.nlm.nih.gov/pubmed/15939837

31. Caspi A, Houts RM, Belsky DW, Harrington H, Hogan S, Ramrakha S, et al. Childhood forecasting of a small segment of the population with large economic burden. Nat Hum Behav [Internet]. 2016 Dec 12;1(1):0005. Available from: https://www.nature.com/articles/s41562-016-0005

32. DiBello JR, Xiong X, Liu X, Zhong W, Merola A, Li M, et al. Trajectories of pharmacological therapies for treatment-resistant depression: a longitudinal study. BMC Psychiatry [Internet]. 2025 Mar 10;25(1):215. Available from:https://bmcpsychiatry.biomedcentral.com/articles/10.1186/s12888-025-06518-8

33. National Institute for Health and Care Excellence (NICE). National Institute for Health and Care Excellence (NICE) Lithium [Internet]. 2025. Available from:https://cks.nice.org.uk/topics/bipolar-disorder/prescribing-information/lithium/#monitoring

34. Cuijpers P, Noma H, Karyotaki E, Vinkers CH, Cipriani A, Furukawa TA. A network meta-analysis of the effects of psychotherapies, pharmacotherapies and their combination in the treatment of adult depression. World Psychiatry [Internet]. 2020 Feb 10;19(1):92–107. Available from: https://onlinelibrary.wiley.com/doi/10.1002/wps.20701

35. Markowitz JC, Wright JH, Peeters F, Thase ME, Kocsis JH, Sudak DM. The Neglected Role of Psychotherapy for Treatment-Resistant Depression. Am J Psychiatry [Internet]. 2022 Feb;179(2):90–3. Available from:https://psychiatryonline.org/doi/10.1176/appi.ajp.2021.21050535

36. Fava M, Rush AJ, Alpert JE, Balasubramani GK, Wisniewski SR, Carmin CN, et al. Difference in Treatment Outcome in Outpatients With Anxious Versus Nonanxious Depression: A STAR*D Report. Am J Psychiatry [Internet]. 2008 Mar;165(3):342–51. Available from: https://psychiatryonline.org/doi/10.1176/appi.ajp.2007.06111868

37. Public Health England. Better care for people with co-occurring mental health and alcohol/drug use conditions A guide for commissioners and service providers [Internet]. 2017. Available from:https://assets.publishing.service.gov.uk/media/5a75b781ed915d6faf2b5276/Co-occurring_mental_health_and_alcohol_drug_use_conditions.pdf

38. Fabbri C, Hagenaars SP, John C, Williams AT, Shrine N, Moles L, et al. Genetic and clinical characteristics of treatment-resistant depression using primary care records in two UK cohorts. Mol Psychiatry [Internet]. 2021 Jul 22;26(7):3363–73. Available from: https://www.nature.com/articles/s41380-021-01062-9

39. Jakubovski E, Bloch MH. Prognostic Subgroups for Citalopram Response in the STAR*D Trial. J Clin Psychiatry [Internet]. 2014 Jul 15;75(07):738–47. Available from: https://www.psychiatrist.com/jcp/prognostic-subgroups-citalopram-response-stard-trial

40. Berlim MT, van den Eynde F, Tovar-Perdomo S, Daskalakis ZJ. Response, remission and drop-out rates following high-frequency repetitive transcranial magnetic stimulation (rTMS) for treating major depression: a systematic review and meta-analysis of randomized, double-blind and sham-controlled trials. Psychol Med [Internet]. 2014 Jan 18;44(2):225–39. Available from: https://www.cambridge.org/core/product/identifier/S0033291713000512/type/journal_article

41. Anand A, Mathew SJ, Sanacora G, Murrough JW, Goes FS, Altinay M, et al. Ketamine versus ECT for Nonpsychotic Treatment-Resistant Major Depression. N Engl J Med [Internet]. 2023 Jun 22;388(25):2315–25. Available from: http://www.nejm.org/doi/10.1056/NEJMoa2302399

42. Carhart-Harris RL, Bolstridge M, Rucker J, Day CMJ, Erritzoe D, Kaelen M, et al. Psilocybin with psychological support for treatment-resistant depression: an open-label feasibility study. The Lancet Psychiatry [Internet]. 2016 Jul;3(7):619–27. Available from: https://linkinghub.elsevier.com/retrieve/pii/S2215036616300657

43. Hannah LA, Walsh CM, Jopling L, Perez J, Cardinal RN, Cameron RA. Economic evaluation of interventions for treatment-resistant depression : A systematic review. 2023;(February):1–20.

