## Supplementary Materials for "Characterising the depression pathway in secondary care: a UK-based epidemiological study of patient characteristics, comorbidities, and treatments"

### SUPPLEMENTARY METHODS

#### Ethical approvals

The Cambridgeshire and Peterborough NHS Foundation Trust (CPFT) Research Database holds ethical approvals from the UK NHS National Research Ethics Service (references 12/EE/0407, 17/EE/0442, 22/EE/0264). Individual consent was neither required nor obtained. The current study was approved by the CPFT Research Database Oversight Committee (local reference Z0009).

#### Eligibility criteria – Diagnosis

Eligible patients included those referred to CPFT with a diagnosis, coded by a CPFT clinician according to the International Classification of Diseases (ICD-10) (25), of F32 (depressive episode) or F33 (recurrent depressive disorder), or the following diagnostic categories also involving the syndrome of depression: F20.4 post-schizophrenic depression; F25.1 schizoaffective disorder, depressive type; F31.3 bipolar affective disorder (BPAD), current episode mild/moderate depression; F31.4 BPAD, severe depression without psychotic symptoms; F31.5 BPAD, severe depression with psychotic symptoms; F38.1 recurrent brief depressive disorder; and F41.2 mixed anxiety and depressive disorder. This included F33 (recurrent depressive disorder) broadly; only 1% (98/9803) of patients had depression coded as in remission (F33.4).

#### Episode of care

The method of establishing the relevant episode of care is shown in **Figure 1**. For the majority of cases, the episode of care spanned the diagnosis (see **Results; Table 1**); however, for a few patients the diagnosis closely preceded the referral, or was recorded shortly after discharge (following a referral starting ≤1 year prior to diagnosis) (**Figure 1**).

Note that this approach does not preclude a different diagnosis before the diagnosis of depression; nor does it exclude the possibility of a previous (e.g. paper-only) referral prior to the introduction of this electronic patient record (EPR) system. This inclusion criterion represents a close electronic approximation to referred patients for whom depression was the primary problem or a major problem with secondary care mental health services. It is possible that different services have varying diagnostic thresholds, leading to an unmeasured diagnosis bias.

#### Clinical directorates

Patients were grouped into age categories according to CPFT's clinical Directorates, according to age at referral: child and adolescent (children and young people's) mental health services (CAMHS) (0–16 years old, inclusive); working-age adults (17–64 years), and older adults (65 years and over).

#### Ethnicity variable

Ethnicity was classified using Office for National Statistics (ONS) categories: White, Asian/Asian British ("Asian"), Black/African/Caribbean/Black British ("Black"), Mixed/Multiple ethnic groups ("Mixed"), Other, and Unknown (1).

#### IMD Variable

The IMD ranks English geographical areas on a scale from 1 (most deprived) to 32,844 (least deprived), incorporating socio-economic factors such as income, employment, education, health, crime, housing and service accessibility, and environmental quality. To use IMD as a continuous predictor, IMD fraction was calculated based on the England range (resulting fraction 0–1, 0 being most deprived). For categorical analyses, IMDs were grouped into quartiles based on the England range (Q1 most deprived, Q4 least deprived).

#### Treatments

**Data pertaining to ECT, inpatient admissions (excluding daycare admissions), and referrals to crisis response/home treatment teams (CRHTTs) were identified using structured referral information. Psychological therapy in secondary care was identified using a structured field classifying the type of progress notes. Data on psychological therapy in primary care was not available.**

#### Gender coding

For a very few patients (< 10; exact number suppressed for statistical disclosure control purposes) gender was reported as either "Other" or "Unknown"; because of these very low numbers, these patients were removed from the dataset and gender was classified as a two-factor variable (male verses female).

#### Medication list

Drugs were grouped into the following categories: selective serotonin reuptake inhibitors (SSRIs) (citalopram, escitalopram, fluoxetine, fluvoxamine, paroxetine, sertraline); serotonin and norepinephrine reuptake inhibitors (SNRIs) (duloxetine, venlafaxine); tricyclics (TCAs) (amitriptyline, clomipramine, dosulepin, doxepin, imipramine, lofepramine, nortriptyline, trimipramine); monoamine oxidase inhibitors (MAOIs) (phenelzine, isocarboxazid, tranylcypromine, moclobemide); mirtazapine; trazodone; second-generation antipsychotics used for antidepressant augmentation according to NICE recommendations (2) (aripiprazole, olanzapine, quetiapine, risperidone); flupentixol (which has a specific license for depression) (3); lithium, lamotrigine, and triiodothyronine (all per NICE guidelines as above). A further category, "Antidepressant broadly, including SGA", captures a union of these drugs. Vortioxetine was not recommended by NICE until after our study end date, so was omitted.

#### Survival analysis

Treatments were analysed separately. For each patient, the analyses began on the date of referral to secondary care.

Some patients were referred to and discharged from CPFT on the same day; for them, the end of observation occurred on the start date, so they were excluded from this analysis.

For some patients, treatment initiation occurred prior to or at the point of referral (e.g. an SSRI documented in a referral letter from primary care). Such patients were excluded from that analysis; thus, each survival analysis describes the rate of initiation of a given treatment for patients with no available documented mention of that treatment before the relevant CPFT referral commenced.

#### References


1. Office for National Statistics. Ethnic group classifications: Census 2021 [Internet]. 2023. Available from: https://www.ons.gov.uk/census/census2021dictionary/variablesbytopic/ethnicgroupnationalidentitylanguageandreligionvariablescensus2021/ethnicgroup/classifications
2. National Institute for Health and Care Excellence. Depression in adults: further-line treatment. 2022;2022. Available from: https://www.nice.org.uk/guidance/ng222/resources/furtherline-treatment-pdf-11131007009
3. Electronic medicines compendium. Electronic medicines compendium - Fluanxol [Internet]. 2021. Available from: https://www.medicines.org.uk/emc/product/11768/smpc


### SUPPLEMENTARY RESULTS

#### Demographics

The mean time from the recording of a depressive disorder to the recording of a psychiatric comorbidity was +23.3 days (comorbidity later), but varied by comorbidity type (**Supplementary Table 1**). However, across all comorbid diagnoses the median time was 0, indicating concurrently coded diagnoses (**Supplementary Table**

**1**); of course, in some cases the comorbidity may have been inherent in the type of depressive disorder (e.g. coding of bipolar affective disorder [BPAD], currently depressed, inherently implies the comorbidity of BPAD). Personality disorders took the longest to diagnose after a depression diagnosis, averaging 161 days (**Supplementary Table 1**).

**Treatment initiation**

The initiation rate for crisis response/home treatment teams (CRHTTs) was closely followed by a parallel curve for inpatient treatment **(Figure 2)**, with approximately 25% of patients utilising these services over time.

**Predictors of treatment**

Patients with substance misuse, psychotic disorders, personality disorders, and self-harm had higher rates of CRHTT referral and inpatient admission (**Supplementary Table 4**). The CAMH group and those with comorbid eating disorders also had higher admission rates. Antipsychotics was initiated more often among patients with comorbid psychotic disorders and BPAD, as might be expected. Furthermore, and as expected, those with BPAD were more commonly prescribed lamotrigine and lithium, and less likely to receive selective serotonin reuptake inhibitor (SSRIs), mirtazapine, serotonin/noradrenaline reuptake inhibitors (SNRIs), and tricyclic antidepressants (TCAs). Patients with comorbid anxiety disorders had significantly lower rates of mirtazapine and lithium use. Psychological therapy in secondary care was more prevalent among those with comorbid anxiety disorders, personality disorders, and eating disorders; and less common in those with organic mental disorders, intellectual disability, or childhood behavioural/emotional disorders. No differences were found in the use of electroconvulsive therapy (ECT), monoamine oxidase inhibitors MAOIs, and triiodothyronine, though this may reflect a lack of power as these treatments were used infrequently.

See also **Supplementary Tables 5–20** for uncorrected analyses contributing to **Supplementary Table 4**.

**Predictors of change**

Individuals with intellectual disabilities improved less than the average, as judged by total Health of the Nation Outcome Scales (HoNOS) scores, while patients with psychotic disorders showed greater overall improvement, but neither were related specifically to the depression sub-score (**Table 2**).

**Sensitivity analysis**

For some patients with BPAD treatment may have been modified to address different symptoms, to account for this we performed a sensitivity analysis excluding BPAD. The effects of MAOIs and lithium were not upheld. In the original analysis we tested for each predictor over and above all other variables including BPAD, therefore this result may have been due to a reduction in power. Alternatively, lithium may be more effective in treating BPAD, and future studies are needed to test this further.

### SUPPLEMENTARY TABLES

#### Supplementary Table 1

***Time of first recording of comorbid diagnoses relative to that of depression.***

| Comorbidity | Mean time from diagnosis of depression to comorbidity (days) | Median (days) |
|---|---|---|
| Organic mental disorders (F0) | 141 (SD 583) | 0 (range −3,701 to 2,686) |
| Substance misuse (F1) | 17.7 (SD 474) | 0 (range −3,150 to 2,395) |
| Psychotic disorders (F2) | −26.4 (SD 1,340) | 0 (range −18,568 to 2,427) |
| Manic episode and bipolar disorder (F30–F31) | −42.9 (SD 856) | 0 (range −13,825 to 2,523) |
| Anxiety disorders and stress reactions (F40–F43) | 41.4 (SD 463) | 0 (range −8,347 to 2,634) |
| Eating disorders (F50) | 2.94 (SD 580) | 0 (range −2,585 to 2,331) |
| Personality disorders (F60–F61) | 161 (SD 654) | 0 (range −4,749 to 2,884) |
| Intellectual disability (F7) | 8.33 (SD 307) | 0 (range −749 to 931) |
| Developmental disorders (F8) | 67.5 (SD 979) | 0 (range −8,766 to 2,457) |
| Childhood behavioural and emotional disorders (F9) | 58.0 (SD 467) | 0 (range −1,400 to 1,626) |
| Any psychiatric comorbidity (F, excluding depression as F20.4, F25.1, F31.3–F31.5, F32–F33, F38.1, F41.2) | 23.3 (SD 719) | 0 (range −18,568 to 2,686) |
| Intentional self-harm (X60–X84) | 159 (SD 562) | 0 (range −1,673 to 2,395) |

#### Supplementary Table 2

***Predictors of duration with CPFT services for index episode (log10-transformed days), including lifetime psychiatric comorbidities.***

| Term | $F$ (via Type III sums of squares) | $p_F$ | Level | Coefficient (95% CI) | Standard error | $t$ | $p_t$ |
|---|---|---|---|---|---|---|---|
| (Intercept) | | | – | +2.16 (CI +2.12 to +2.21) | 0.0242 | $t_{8937}$ = +89.5 | $p < 2.20 \times 10^{-16}$ ***** |
| Service (age band) at referral | $F_{2,8937} = 63.8$ | $p < 2.20 \times 10^{-16}$ ***** | Adult | Reference | | | |
| | | | CAMHS | +0.444 (CI +0.345 to +0.543) | 0.0505 | $t_{8937}$ = +8.79 | $p < 2.20 \times 10^{-16}$ ***** |
| | | | Older adult | −0.119 (CI −0.156 to −0.0811) | 0.0191 | $t_{8937}$ = −6.20 | $p = 6.04 \times 10^{-10}$ ***** |
| Sex | $F_{1,8937} = 49.9$ | $p = 1.73 \times 10^{-12}$ ***** | Female | Reference | | | |
| | | | Male | −0.116 (CI −0.148 to −0.0838) | 0.0164 | $t_{8937}$ = −7.06 | $p = 1.73 \times 10^{-12}$ ***** |
| Ethnicity | $F_{5,8937} = 39.7$ | $p < 2.20 \times 10^{-16}$ ***** | White | Reference | | | |
| | | | Asian | +0.00403 (CI −0.103 to +0.111) | 0.0546 | $\|t\| < 1$ | NS |

| Term | $F$ (via Type III sums of squares) | $p_F$ | Level | Coefficient (95% CI) | Standard error | $t$ | $p_t$ |
|---|---|---|---|---|---|---|---|
| | | | Black | −0.0192 (CI −0.200 to +0.162) | 0.0922 | $\|t\| < 1$ | NS |
| | | | Mixed | +0.00646 (CI −0.137 to +0.150) | 0.0731 | $\|t\| < 1$ | NS |
| | | | Other | +0.0280 (CI −0.125 to +0.181) | 0.0780 | $\|t\| < 1$ | NS |
| | | | Unknown | −0.306 (CI −0.349 to −0.263) | 0.0218 | $t_{8937} = -14.0$ | $p < 2.20 \times 10^{-16}$ ***** |
| IMD fraction (0–1, higher values indicating less deprived) | $F_{1,8937} = 7.62$ | $p = 0.00580$ ** | – | −0.0860 (CI −0.147 to −0.0249) | 0.0312 | $t_{8937} = -2.76$ | $p = 0.00580$ ** |
| Organic mental disorders (F0) | $F < 1$ | NS | | | | | |
| Substance misuse (F1) | $F_{1,8937} = 20.0$ | $p = 7.90 \times 10^{-6}$ ***** | True | −0.145 (CI −0.208 to −0.0813) | 0.0324 | $t_{8937} = -4.47$ | $p = 7.90 \times 10^{-6}$ ***** |
| Psychotic disorders (F2) | $F_{1,8937} = 64.5$ | $p = 1.09 \times 10^{-15}$ ***** | True | +0.362 (CI +0.274 to +0.451) | 0.0451 | $t_{8937} = +8.03$ | $p = 1.09 \times 10^{-15}$ ***** |
| Manic episode and bipolar disorder (F30–F31) | $F_{1,8937} = 146$ | $p < 2.20 \times 10^{-16}$ ***** | True | +0.399 (CI +0.334 to +0.463) | 0.0330 | $t_{8937} = +12.1$ | $p < 2.20 \times 10^{-16}$ ***** |
| Anxiety disorders and stress reactions (F40–F43) | $F_{1,8937} = 85.3$ | $p < 2.20 \times 10^{-16}$ ***** | True | +0.163 (CI +0.128 to +0.197) | 0.0176 | $t_{8937} = +9.24$ | $p < 2.20 \times 10^{-16}$ ***** |
| Eating disorders (F50) | $F_{1,8937} = 13.9$ | $p = 0.000196$ *** | True | +0.187 (CI +0.0886 to +0.285) | 0.0502 | $t_{8937} = +3.73$ | $p = 0.000196$ *** |
| Personality disorders (F60–F61) | $F_{1,8937} = 47.1$ | $p = 7.32 \times 10^{-12}$ ***** | True | +0.206 (CI +0.147 to +0.264) | 0.0300 | $t_{8937} = +6.86$ | $p = 7.32 \times 10^{-12}$ ***** |
| Intellectual disability (F7) | $F_{1,8937} = 29.6$ | $p = 5.32 \times 10^{-8}$ ***** | True | +0.645 (CI +0.413 to +0.877) | 0.118 | $t_{8937} = +5.45$ | $p = 5.32 \times 10^{-8}$ ***** |
| Developmental disorders (F8) | $F_{1,8937} = 18.2$ | $p = 2.01 \times 10^{-5}$ **** | True | +0.275 (CI +0.149 to +0.402) | 0.0645 | $t_{8937} = +4.27$ | $p = 2.01 \times 10^{-5}$ **** |
| Childhood behavioural and emotional disorders (F9) | $F_{1,8937} = 11.9$ | $p = 0.000555$ *** | True | +0.213 (CI +0.0923 to +0.335) | 0.0618 | $t_{8937} = +3.45$ | $p = 0.000555$ *** |
| Intentional self-harm (X60–X84) | $F_{1,8937} = 65.6$ | $p = 6.40 \times 10^{-16}$ ***** | True | −0.314 (CI −0.389 to −0.238) | 0.0387 | $t_{8937} = -8.10$ | $p = 6.40 \times 10^{-16}$ ***** |

### Supplementary Table 3

***Predictors of first relevant HoNOS scores (lower values are better), for all those with at least one time-relevant HoNOS score.***

| Term | $F$ (via Type III sums of squares) | $p_F$ | Level | Coefficient (95% CI) | Standard error | $t$ | $p_t$ |
|---|---|---|---|---|---|---|---|
| **DEPRESSION SUB-SCORES** | | | | | | | |
| (Intercept) | | | – | +2.53 (CI +2.47 to +2.60) | 0.0334 | $t_{6025} = +75.9$ | $p < 2.20 \times 10^{-16}$ ***** |
| Service (age band) at referral | $F_{2,6025} = 33.8$ | $p = 2.41 \times 10^{-15}$ ***** | Adult | Reference | | | |
| | | | CAMHS | −0.310 (CI −0.574 to −0.0470) | 0.134 | $t_{6025} = -2.31$ | $p = 0.0209$ * |
| | | | Older adult | −0.216 (CI −0.269 to −0.163) | 0.0270 | $t_{6025} = -7.98$ | $p = 1.68 \times 10^{-15}$ ***** |
| Sex | $F_{1,6025} = 1.91$ | $p = 0.167$, NS | | | | | |

| Term | $F$ (via Type III sums of squares) | $p_F$ | Level | Coefficient (95% CI) | Standard error | $t$ | $p_t$ |
|---|---|---|---|---|---|---|---|
| Ethnicity | $F_{5,6025} = 6.30$ | $p = 7.75 \times 10^{-6}$ ***** | White | Reference | | | |
| | | | Asian | +0.0904 (CI −0.0571 to +0.238) | 0.0752 | $t_{6025}$ = +1.20 | $p = 0.229$, NS |
| | | | Black | −0.0323 (CI −0.273 to +0.209) | 0.123 | $\|t\| < 1$ | NS |
| | | | Mixed | −0.0218 (CI −0.217 to +0.173) | 0.0994 | $\|t\| < 1$ | NS |
| | | | Other | +0.0608 (CI −0.148 to +0.269) | 0.106 | $\|t\| < 1$ | NS |
| | | | Unknown | −0.188 (CI −0.258 to −0.119) | 0.0353 | $t_{6025}$ = −5.34 | $p = 9.73 \times 10^{-8}$ ***** |
| IMD fraction (0–1, higher values indicating less deprived) | $F_{1,6025} = 2.21$ | $p = 0.138$, NS | | | | | |
| Organic mental disorders (F0) | $F_{1,6025} = 176$ | $p < 2.20 \times 10^{-16}$ ***** | True | −0.491 (CI −0.564 to −0.419) | 0.0370 | $t_{6025}$ = −13.3 | $p < 2.20 \times 10^{-16}$ ***** |
| Substance misuse (F1) | $F_{1,6025} = 24.4$ | $p = 7.96 \times 10^{-7}$ ***** | True | −0.217 (CI −0.303 to −0.131) | 0.0438 | $t_{6025}$ = −4.94 | $p = 7.96 \times 10^{-7}$ ***** |
| Psychotic disorders (F2) | $F_{1,6025} = 92.6$ | $p < 2.20 \times 10^{-16}$ ***** | True | −0.556 (CI −0.669 to −0.443) | 0.0578 | $t_{6025}$ = −9.62 | $p < 2.20 \times 10^{-16}$ ***** |
| Manic episode and bipolar disorder (F30–F31) | $F_{1,6025} = 48.5$ | $p = 3.73 \times 10^{-12}$ ***** | True | −0.292 (CI −0.375 to −0.210) | 0.0420 | $t_{6025}$ = −6.96 | $p = 3.73 \times 10^{-12}$ ***** |
| Anxiety disorders and stress reactions (F40–F43) | $F_{1,6025} = 19.1$ | $p = 1.29 \times 10^{-5}$ **** | True | −0.105 (CI −0.153 to −0.0581) | 0.0241 | $t_{6025}$ = −4.37 | $p = 1.29 \times 10^{-5}$ **** |
| Eating disorders (F50) | $F_{1,6025} = 1.07$ | $p = 0.300$, NS | | | | | |
| Personality disorders (F60–F61) | $F_{1,6025} = 5.36$ | $p = 0.0206$ * | True | −0.0903 (CI −0.167 to −0.0138) | 0.0390 | $t_{6025}$ = −2.32 | $p = 0.0206$ * |
| Intellectual disability (F7) | $F_{1,6025} = 2.34$ | $p = 0.126$, NS | | | | | |
| Developmental disorders (F8) | $F_{1,6025} = 3.39$ | $p = 0.0655$, NS | | | | | |
| Childhood behavioural and emotional disorders (F9) | $F_{1,6025} = 1.03$ | $p = 0.311$, NS | | | | | |
| Intentional self-harm (X60–X84) | $F < 1$ | NS | | | | | |
| **TOTAL SCORES** | | | | | | | |
| (Intercept) | | | – | +13.0 (CI +12.6 to +13.4) | 0.189 | $t_{6025}$ = +69.0 | $p < 2.20 \times 10^{-16}$ ***** |
| Service (age band) at referral | $F_{2,6025} = 63.2$ | $p < 2.20 \times 10^{-16}$ ***** | Adult | Reference | | | |
| | | | CAMHS | −0.313 (CI −1.80 to +1.17) | 0.759 | $\|t\| < 1$ | NS |
| | | | Older adult | −1.72 (CI −2.02 to −1.42) | 0.153 | $t_{6025}$ = −11.2 | $p < 2.20 \times 10^{-16}$ ***** |
| Sex | $F_{1,6025} = 28.5$ | $p = 9.71 \times 10^{-8}$ ***** | Female | Reference | | | |
| | | | Male | +0.702 (CI +0.444 to +0.960) | 0.131 | $t_{6025}$ = +5.34 | $p = 9.71 \times 10^{-8}$ ***** |

| Term | *F* (via Type III sums of squares) | $p_F$ | Level | Coefficient (95% CI) | Standard error | *t* | $p_t$ |
|---|---|---|---|---|---|---|---|
| Ethnicity | $F_{5,6025} = 8.81$ | $p = 2.41 \times 10^{-8}$ ***** | White | Reference | | | |
| | | | Asian | +0.543 (CI −0.290 to +1.38) | 0.425 | $t_{6025} = +1.28$ | $p = 0.201$, NS |
| | | | Black | +0.900 (CI −0.462 to +2.26) | 0.695 | $t_{6025} = +1.30$ | $p = 0.195$, NS |
| | | | Mixed | −0.836 (CI −1.94 to +0.265) | 0.562 | $t_{6025} = -1.49$ | $p = 0.137$, NS |
| | | | Other | +0.158 (CI −1.02 to +1.34) | 0.601 | $\|t\| < 1$ | NS |
| | | | Unknown | −1.22 (CI −1.61 to −0.826) | 0.199 | $t_{6025} = -6.10$ | $p = 1.11 \times 10^{-9}$ ***** |
| IMD fraction (0–1, higher values indicating less deprived) | $F_{1,6025} = 37.1$ | $p = 1.21 \times 10^{-9}$ ***** | – | −1.49 (CI −1.97 to −1.01) | 0.244 | $t_{6025} = -6.09$ | $p = 1.21 \times 10^{-9}$ ***** |
| Organic mental disorders (F0) | $F_{1,6025} = 1.32$ | $p = 0.250$, NS | | | | | |
| Substance misuse (F1) | $F_{1,6025} = 7.19$ | $p = 0.00735$ ** | True | +0.664 (CI +0.179 to +1.15) | 0.248 | $t_{6025} = +2.68$ | $p = 0.00735$ ** |
| Psychotic disorders (F2) | $F_{1,6025} = 3.82$ | $p = 0.0506$, NS | | | | | |
| Manic episode and bipolar disorder (F30–F31) | $F_{1,6025} = 28.0$ | $p = 1.28 \times 10^{-7}$ ***** | True | −1.25 (CI −1.72 to −0.790) | 0.237 | $t_{6025} = -5.29$ | $p = 1.28 \times 10^{-7}$ ***** |
| Anxiety disorders and stress reactions (F40–F43) | $F < 1$ | NS | | | | | |
| Eating disorders (F50) | $F_{1,6025} = 91.9$ | $p < 2.20 \times 10^{-16}$ ***** | True | +4.17 (CI +3.32 to +5.03) | 0.435 | $t_{6025} = +9.59$ | $p < 2.20 \times 10^{-16}$ ***** |
| Personality disorders (F60–F61) | $F_{1,6025} = 9.57$ | $p = 0.00198$ ** | True | +0.682 (CI +0.250 to +1.11) | 0.220 | $t_{6025} = +3.09$ | $p = 0.00198$ ** |
| Intellectual disability (F7) | $F < 1$ | NS | | | | | |
| Developmental disorders (F8) | $F_{1,6025} = 8.53$ | $p = 0.00351$ ** | True | +1.49 (CI +0.490 to +2.49) | 0.511 | $t_{6025} = +2.92$ | $p = 0.00351$ ** |
| Childhood behavioural and emotional disorders (F9) | $F < 1$ | NS | | | | | |
| Intentional self-harm (X60–X84) | $F < 1$ | NS | | | | | |

### Supplementary Table 4

Summary of predictors of treatments. Each column represents a single Cox proportional hazards model for a single treatment type (see following tables for each model separately), but here Šidák corrections are applied to *α* for a family of 16 models. (ADep, antidepressants as a broad category. Adm, inpatient admission to mental health hospital. APsy, NICE [United Kingdom National Institute for Health and Care Excellence]-endorsed antipsychotics for antidepressant augmentation. CAMHS, child and adolescent mental health services. Crisis, referral to crisis response/home treatment team. ECT, electroconvulsive therapy.Flu, flupentixol. IMD, Index of Multiple Deprivation. Lam, lamotrigine. Li, lithium. MAOI, monoamine oxidase inhibitor. Mirt, mirtazapine. Psychol, psychology service in secondary care. SNRI, serotonin/noradrenaline reuptake inhibitor. SSRI, selective serotonin reuptake inhibitor. T3, triiodothyronine. TCA, tricyclic antidepressant. Traz, trazodone.)

| Term | Level | ADep | SSRI | Mirt | APsy | SNRI | TCA | Lam | Li | Traz | MAOI | Flu | T3 | Psychol | Crisis | Adm | ECT |
|---|---|---|---|---|---|---|---|---|---|---|---|---|---|---|---|---|---|
| Service (age band) at referral | Adult | | | | | | | | | | | | | | | | |
| | CAMHS | ↓ | ↔ | ↓ | ↓ | ↓ | ↓ | ↔ | ↔ | ↔ | ↔ | ↔ | ↔ | ↑ | ↓ | ↑ | ↔ |
| | Older adult | ↓ | ↓ | ↑ | ↓ | ↓ | ↑ | ↓ | ↔ | ↔ | ↔ | ↔ | ↔ | ↑ | ↔ | ↓ | ↔ |
| Sex | Female | | | | | | | | | | | | | | | | |

| Term | Level | ADep | SSRI | Mirt | APsy | SNRI | TCA | Lam | Li | Traz | MAOI | Flu | T3 | Psychol | Crisis | Adm | ECT |
|---|---|---|---|---|---|---|---|---|---|---|---|---|---|---|---|---|---|
| | Male | ↔ | ↔ | ↑ | ↔ | ↔ | ↓ | ↔ | ↔ | ↔ | ↔ | ↔ | ↔ | ↔ | ↔ | ↑ | ↔ |
| Ethnicity | White | | | | | | | | | | | | | | | | |
| | Asian | ↔ | ↔ | ↔ | ↔ | ↔ | ↔ | ↔ | ↔ | ↔ | ↔ | ↔ | ↔ | ↔ | ↔ | ↔ | ↔ |
| | Black | ↔ | ↔ | ↔ | ↔ | ↔ | ↔ | ↔ | ↔ | ↔ | ↔ | ↔ | ↔ | ↔ | ↔ | ↔ | ↔ |
| | Mixed | ↔ | ↔ | ↔ | ↔ | ↔ | ↔ | ↔ | ↔ | ↑ | ↔ | ↔ | ↔ | ↔ | ↔ | ↔ | ↔ |
| | Other | ↔ | ↔ | ↔ | ↔ | ↔ | ↔ | ↔ | ↔ | ↔ | ↔ | ↔ | ↔ | ↔ | ↔ | ↔ | ↔ |
| | Unknown | ↔ | ↔ | ↔ | ↓ | ↔ | ↔ | ↔ | ↓ | ↔ | ↔ | ↔ | ↔ | ↓ | ↓ | ↓ | ↔ |
| IMD fraction (0–1, higher values indicating less deprived) | – | ↔ | ↔ | ↔ | ↔ | ↔ | ↔ | ↔ | ↑ | ↔ | ↔ | ↔ | ↔ | ↔ | ↔ | ↓ | ↔ |
| Organic mental disorders (F0) | True | ↓ | ↔ | ↔ | ↔ | ↓ | ↔ | ↔ | ↔ | ↔ | ↔ | ↔ | ↔ | ↓ | ↔ | ↔ | ↔ |
| Substance misuse (F1) | True | ↔ | ↔ | ↑ | ↔ | ↔ | ↔ | ↔ | ↔ | ↔ | ↔ | ↔ | ↔ | ↔ | ↑ | ↑ | ↔ |
| Psychotic disorders (F2) | True | ↔ | ↔ | ↔ | ↑ | ↔ | ↔ | ↔ | ↔ | ↔ | ↔ | ↑ | ↔ | ↔ | ↑ | ↑ | ↔ |
| Manic episode and bipolar disorder (F30–F31) | True | ↔ | ↓ | ↓ | ↑ | ↓ | ↓ | ↑ | ↑ | ↔ | ↔ | ↔ | ↔ | ↔ | ↔ | ↔ | ↔ |
| Anxiety disorders and stress reactions (F40–F43) | True | ↓ | ↔ | ↓ | ↔ | ↔ | ↔ | ↔ | ↓ | ↔ | ↔ | ↔ | ↔ | ↑ | ↔ | ↔ | ↔ |
| Eating disorders (F50) | True | ↔ | ↔ | ↔ | ↔ | ↔ | ↔ | ↔ | ↔ | ↔ | ↔ | ↔ | ↔ | ↑ | ↔ | ↑ | ↔ |
| Personality disorders (F60–F61) | True | ↔ | ↔ | ↔ | ↑ | ↑ | ↔ | ↔ | ↔ | ↑ | ↔ | ↔ | ↔ | ↑ | ↑ | ↑ | ↔ |
| Intellectual disability (F7) | True | ↓ | ↔ | ↓ | ↔ | ↓ | ↔ | ↔ | ↔ | ↔ | ↔ | ↔ | ↔ | ↓ | ↔ | ↔ | ↔ |
| Developmental disorders (F8) | True | ↔ | ↔ | ↔ | ↔ | ↔ | ↔ | ↔ | ↔ | ↔ | ↔ | ↔ | ↔ | ↔ | ↔ | ↔ | ↔ |
| Childhood behavioural and emotional disorders (F9) | True | ↓ | ↔ | ↔ | ↔ | ↓ | ↔ | ↔ | ↔ | ↔ | ↔ | ↔ | ↔ | ↓ | ↔ | ↔ | ↔ |
| Intentional self-harm (X60–X84) | True | ↑ | ↑ | ↔ | ↔ | ↔ | ↔ | ↔ | ↔ | ↔ | ↔ | ↔ | ↔ | ↔ | ↑ | ↑ | ↔ |

### Supplementary Table 5

***Predictors of antidepressant use (broadly defined inc. SGAs)***

| Term | Level | Coefficient | $e^{coeff}$ | SE(coeff) | $Z$ | $p_{\|Z\|}$ |
|---|---|---|---|---|---|---|
| Service (age band) at referral | Adult | Reference | | | | |
| | CAMHS | −0.825 | 0.438 (CI 0.380 to 0.506) | 0.0733 | −11.3 | $p < 2.20 \times 10^{-16}$ ***** |
| | Older adult | −0.115 | 0.891 (CI 0.838 to 0.948) | 0.0314 | −3.67 | $p = 0.000242$ *** |
| Sex | Female | Reference | | | | |
| | Male | +0.0725 | 1.08 (CI 1.02 to 1.13) | 0.0271 | +2.68 | $p = 0.00747$ ** |

| Term | Level | Coefficient | $e^{coeff}$ | SE(coeff) | $Z$ | $p_{\|Z\|}$ |
|---|---|---|---|---|---|---|
| Ethnicity | White | Reference | | | | |
| | Asian | +0.0586 | 1.06 (CI 0.895 to 1.26) | 0.0866 | +0.676 | $p$ = 0.499, NS |
| | Black | +0.122 | 1.13 (CI 0.852 to 1.50) | 0.144 | +0.848 | $p$ = 0.396, NS |
| | Mixed | +0.161 | 1.17 (CI 0.938 to 1.47) | 0.115 | +1.41 | $p$ = 0.160, NS |
| | Other | −0.151 | 0.860 (CI 0.663 to 1.12) | 0.133 | −1.14 | $p$ = 0.255, NS |
| | Unknown | −0.0439 | 0.957 (CI 0.890 to 1.03) | 0.0371 | −1.18 | $p$ = 0.236, NS |
| IMD fraction (0–1, higher values indicating less deprived) | – | +0.108 | 1.11 (CI 1.01 to 1.23) | 0.0512 | +2.10 | $p$ = 0.0358 * |
| Organic mental disorders (F0) | True | −0.167 | 0.847 (CI 0.782 to 0.917) | 0.0406 | −4.10 | $p = 4.07 \times 10^{-5}$ **** |
| Substance misuse (F1) | True | +0.0358 | 1.04 (CI 0.932 to 1.15) | 0.0540 | +0.663 | $p$ = 0.508, NS |
| Psychotic disorders (F2) | True | +0.0916 | 1.10 (CI 0.957 to 1.26) | 0.0692 | +1.32 | $p$ = 0.185, NS |
| Manic episode and bipolar disorder (F30–F31) | True | −0.0972 | 0.907 (CI 0.819 to 1.00) | 0.0521 | −1.87 | $p$ = 0.0619, NS |
| Anxiety disorders and stress reactions (F40–F43) | True | −0.145 | 0.865 (CI 0.817 to 0.915) | 0.0290 | −5.01 | $p = 5.44 \times 10^{-7}$ ***** |
| Eating disorders (F50) | True | −0.151 | 0.860 (CI 0.740 to 0.998) | 0.0762 | −1.98 | $p$ = 0.0472 * |
| Personality disorders (F60–F61) | True | +0.0411 | 1.04 (CI 0.951 to 1.14) | 0.0469 | +0.877 | $p$ = 0.380, NS |
| Intellectual disability (F7) | True | −0.529 | 0.589 (CI 0.418 to 0.830) | 0.175 | −3.02 | $p$ = 0.00250 ** |
| Developmental disorders (F8) | True | −0.123 | 0.884 (CI 0.723 to 1.08) | 0.103 | −1.19 | $p$ = 0.232, NS |
| Childhood behavioural and emotional disorders (F9) | True | −0.391 | 0.677 (CI 0.562 to 0.815) | 0.0947 | −4.13 | $p = 3.69 \times 10^{-5}$ **** |
| Intentional self-harm (X60–X84) | True | +0.271 | 1.31 (CI 1.15 to 1.50) | 0.0687 | +3.95 | $p = 7.84 \times 10^{-5}$ **** |

### Supplementary Table 6

***Predictors of SSRI use***

| Term | Level | Coefficient | $e^{coeff}$ | SE(coeff) | $Z$ | $p_{\|Z\|}$ |
|---|---|---|---|---|---|---|
| Service (age band) at referral | Adult | Reference | | | | |
| | CAMHS | −0.123 | 0.884 (CI 0.762 to 1.03) | 0.0756 | −1.63 | $p$ = 0.103, NS |
| | Older adult | −0.320 | 0.726 (CI 0.674 to 0.782) | 0.0377 | −8.51 | $p < 2.20 \times 10^{-16}$ ***** |
| Sex | Female | Reference | | | | |
| | Male | +0.0262 | 1.03 (CI 0.965 to 1.09) | 0.0317 | +0.828 | $p$ = 0.407, NS |
| Ethnicity | White | Reference | | | | |

| Term | Level | Coefficient | $e^{coeff}$ | SE(coeff) | $Z$ | $p_{\|Z\|}$ |
|---|---|---|---|---|---|---|
| | Asian | +0.00899 | 1.01 (CI 0.829 to 1.23) | 0.100 | +0.0896 | $p$ = 0.929, NS |
| | Black | +0.0773 | 1.08 (CI 0.777 to 1.50) | 0.168 | +0.460 | $p$ = 0.646, NS |
| | Mixed | +0.250 | 1.28 (CI 1.00 to 1.65) | 0.127 | +1.98 | $p$ = 0.0482 * |
| | Other | +0.136 | 1.15 (CI 0.881 to 1.49) | 0.134 | +1.01 | $p$ = 0.311, NS |
| | Unknown | +0.0217 | 1.02 (CI 0.937 to 1.11) | 0.0444 | +0.490 | $p$ = 0.624, NS |
| IMD fraction (0–1, higher values indicating less deprived) | – | +0.0357 | 1.04 (CI 0.923 to 1.16) | 0.0591 | +0.604 | $p$ = 0.546, NS |
| Organic mental disorders (F0) | True | −0.0675 | 0.935 (CI 0.848 to 1.03) | 0.0500 | −1.35 | $p$ = 0.177, NS |
| Substance misuse (F1) | True | −0.0190 | 0.981 (CI 0.868 to 1.11) | 0.0627 | −0.304 | $p$ = 0.761, NS |
| Psychotic disorders (F2) | True | −0.179 | 0.836 (CI 0.715 to 0.978) | 0.0798 | −2.24 | $p$ = 0.0252 * |
| Manic episode and bipolar disorder (F30–F31) | True | −0.408 | 0.665 (CI 0.588 to 0.752) | 0.0626 | −6.52 | $p = 6.91 \times 10^{-11}$ ***** |
| Anxiety disorders and stress reactions (F40–F43) | True | −0.0208 | 0.979 (CI 0.918 to 1.04) | 0.0330 | −0.632 | $p$ = 0.528, NS |
| Eating disorders (F50) | True | −0.0224 | 0.978 (CI 0.829 to 1.15) | 0.0840 | −0.267 | $p$ = 0.790, NS |
| Personality disorders (F60–F61) | True | −0.0530 | 0.948 (CI 0.854 to 1.05) | 0.0533 | −0.993 | $p$ = 0.321, NS |
| Intellectual disability (F7) | True | −0.183 | 0.833 (CI 0.573 to 1.21) | 0.191 | −0.957 | $p$ = 0.339, NS |
| Developmental disorders (F8) | True | −0.107 | 0.899 (CI 0.720 to 1.12) | 0.113 | −0.942 | $p$ = 0.346, NS |
| Childhood behavioural and emotional disorders (F9) | True | −0.117 | 0.890 (CI 0.726 to 1.09) | 0.104 | −1.13 | $p$ = 0.260, NS |
| Intentional self-harm (X60–X84) | True | +0.263 | 1.30 (CI 1.12 to 1.51) | 0.0764 | +3.44 | $p$ = 0.000574 *** |

### Supplementary Table 7

***Predictors of mirtazapine use***

| Term | Level | Coefficient | $e^{coeff}$ | SE(coeff) | $Z$ | $p_{\|Z\|}$ |
|---|---|---|---|---|---|---|
| Service (age band) at referral | Adult | Reference | | | | |
| | CAMHS | −0.797 | 0.451 (CI 0.335 to 0.607) | 0.152 | −5.25 | $p = 1.50 \times 10^{-7}$ ***** |
| | Older adult | +0.673 | 1.96 (CI 1.80 to 2.13) | 0.0432 | +15.6 | $p < 2.20 \times 10^{-16}$ ***** |
| Sex | Female | Reference | | | | |
| | Male | +0.153 | 1.17 (CI 1.08 to 1.26) | 0.0377 | +4.07 | $p = 4.69 \times 10^{-5}$ **** |
| Ethnicity | White | Reference | | | | |
| | Asian | −0.0712 | 0.931 (CI 0.718 to 1.21) | 0.133 | −0.537 | $p$ = 0.591, NS |

| Term | Level | Coefficient | $e^{coeff}$ | SE(coeff) | $Z$ | $p_{\|Z\|}$ |
|---|---|---|---|---|---|---|
| | Black | −0.133 | 0.875 (CI 0.557 to 1.38) | 0.231 | −0.576 | $p$ = 0.565, NS |
| | Mixed | −0.438 | 0.646 (CI 0.431 to 0.966) | 0.206 | −2.13 | $p$ = 0.0332 * |
| | Other | +0.0738 | 1.08 (CI 0.750 to 1.55) | 0.185 | +0.400 | $p$ = 0.689, NS |
| | Unknown | −0.0701 | 0.932 (CI 0.838 to 1.04) | 0.0546 | −1.28 | $p$ = 0.199, NS |
| IMD fraction (0–1, higher values indicating less deprived) | – | −0.127 | 0.881 (CI 0.766 to 1.01) | 0.0712 | −1.78 | $p$ = 0.0745, NS |
| Organic mental disorders (F0) | True | +0.0637 | 1.07 (CI 0.964 to 1.18) | 0.0509 | +1.25 | $p$ = 0.212, NS |
| Substance misuse (F1) | True | +0.277 | 1.32 (CI 1.15 to 1.52) | 0.0723 | +3.84 | $p$ = 0.000124 *** |
| Psychotic disorders (F2) | True | −0.244 | 0.783 (CI 0.639 to 0.961) | 0.104 | −2.34 | $p$ = 0.0193 * |
| Manic episode and bipolar disorder (F30–F31) | True | −0.770 | 0.463 (CI 0.387 to 0.554) | 0.0914 | −8.42 | $p < 2.20 \times 10^{-16}$ ***** |
| Anxiety disorders and stress reactions (F40–F43) | True | −0.120 | 0.887 (CI 0.819 to 0.960) | 0.0407 | −2.96 | $p$ = 0.00308 ** |
| Eating disorders (F50) | True | −0.397 | 0.673 (CI 0.512 to 0.883) | 0.139 | −2.85 | $p$ = 0.00433 ** |
| Personality disorders (F60–F61) | True | +0.0459 | 1.05 (CI 0.913 to 1.20) | 0.0701 | +0.655 | $p$ = 0.513, NS |
| Intellectual disability (F7) | True | −1.63 | 0.197 (CI 0.0815 to 0.475) | 0.450 | −3.62 | $p$ = 0.000298 *** |
| Developmental disorders (F8) | True | −0.293 | 0.746 (CI 0.537 to 1.04) | 0.168 | −1.75 | $p$ = 0.0804, NS |
| Childhood behavioural and emotional disorders (F9) | True | −0.447 | 0.640 (CI 0.459 to 0.892) | 0.170 | −2.63 | $p$ = 0.00851 ** |
| Intentional self-harm (X60–X84) | True | +0.0855 | 1.09 (CI 0.904 to 1.31) | 0.0952 | +0.897 | $p$ = 0.369, NS |

### Supplementary Table 8

***Predictors of NICE-endorsed augmentation antipsychotic use***

| Term | Level | Coefficient | $e^{coeff}$ | SE(coeff) | $Z$ | $p_{\|Z\|}$ |
|---|---|---|---|---|---|---|
| Service (age band) at referral | Adult | Reference | | | | |
| | CAMHS | −0.580 | 0.560 (CI 0.442 to 0.709) | 0.120 | −4.82 | $p = 1.46 \times 10^{-6}$ ***** |
| | Older adult | −0.713 | 0.490 (CI 0.439 to 0.547) | 0.0558 | −12.8 | $p < 2.20 \times 10^{-16}$ ***** |
| Sex | Female | Reference | | | | |
| | Male | +0.0153 | 1.02 (CI 0.933 to 1.10) | 0.0429 | +0.357 | $p$ = 0.721, NS |
| Ethnicity | White | Reference | | | | |
| | Asian | +0.0111 | 1.01 (CI 0.788 to 1.30) | 0.127 | +0.0875 | $p$ = 0.930, NS |
| | Black | +0.504 | 1.66 (CI 1.18 to 2.33) | 0.174 | +2.89 | $p$ = 0.00386 ** |
| | Mixed | −0.127 | 0.881 (CI 0.610 to 1.27) | 0.188 | −0.675 | $p$ = 0.500, NS |
| | Other | −0.0474 | 0.954 (CI 0.651 to 1.40) | 0.195 | −0.243 | $p$ = 0.808, NS |

| Term | Level | Coefficient | $e^{coeff}$ | SE(coeff) | $Z$ | $p_{\|Z\|}$ |
|---|---|---|---|---|---|---|
| | Unknown | −0.352 | 0.704 (CI 0.613 to 0.808) | 0.0706 | −4.98 | $p = 6.37 \times 10^{-7}$ ***** |
| IMD fraction (0–1, higher values indicating less deprived) | – | +0.107 | 1.11 (CI 0.953 to 1.30) | 0.0790 | +1.35 | $p$ = 0.177, NS |
| Organic mental disorders (F0) | True | −0.0345 | 0.966 (CI 0.829 to 1.13) | 0.0782 | −0.441 | $p$ = 0.659, NS |
| Substance misuse (F1) | True | +0.181 | 1.20 (CI 1.03 to 1.39) | 0.0751 | +2.40 | $p$ = 0.0162 * |
| Psychotic disorders (F2) | True | +1.32 | 3.76 (CI 3.25 to 4.34) | 0.0734 | +18.0 | $p < 2.20 \times 10^{-16}$ ***** |
| Manic episode and bipolar disorder (F30–F31) | True | +0.816 | 2.26 (CI 2.01 to 2.55) | 0.0608 | +13.4 | $p < 2.20 \times 10^{-16}$ ***** |
| Anxiety disorders and stress reactions (F40–F43) | True | −0.0122 | 0.988 (CI 0.905 to 1.08) | 0.0446 | −0.274 | $p$ = 0.784, NS |
| Eating disorders (F50) | True | −0.0983 | 0.906 (CI 0.732 to 1.12) | 0.109 | −0.899 | $p$ = 0.369, NS |
| Personality disorders (F60–F61) | True | +0.441 | 1.55 (CI 1.39 to 1.74) | 0.0586 | +7.53 | $p = 4.93 \times 10^{-14}$ ***** |
| Intellectual disability (F7) | True | −0.459 | 0.632 (CI 0.372 to 1.08) | 0.271 | −1.69 | $p$ = 0.0908, NS |
| Developmental disorders (F8) | True | −0.121 | 0.886 (CI 0.669 to 1.17) | 0.144 | −0.840 | $p$ = 0.401, NS |
| Childhood behavioural and emotional disorders (F9) | True | −0.358 | 0.699 (CI 0.514 to 0.950) | 0.157 | −2.29 | $p$ = 0.0220 * |
| Intentional self-harm (X60–X84) | True | +0.174 | 1.19 (CI 0.988 to 1.43) | 0.0950 | +1.83 | $p$ = 0.0675, NS |

### Supplementary Table 9

***Predictors of SNRI use***

| Term | Level | Coefficient | $e^{coeff}$ | SE(coeff) | $Z$ | $p_{\|Z\|}$ |
|---|---|---|---|---|---|---|
| Service (age band) at referral | Adult | Reference | | | | |
| | CAMHS | −1.64 | 0.194 (CI 0.124 to 0.303) | 0.227 | −7.22 | $p = 5.02 \times 10^{-13}$ ***** |
| | Older adult | −0.338 | 0.713 (CI 0.635 to 0.801) | 0.0592 | −5.71 | $p = 1.13 \times 10^{-8}$ ***** |
| Sex | Female | Reference | | | | |
| | Male | +0.0102 | 1.01 (CI 0.917 to 1.11) | 0.0496 | +0.206 | $p$ = 0.836, NS |
| Ethnicity | White | Reference | | | | |
| | Asian | −0.226 | 0.798 (CI 0.577 to 1.10) | 0.165 | −1.37 | $p$ = 0.172, NS |
| | Black | −0.399 | 0.671 (CI 0.370 to 1.22) | 0.303 | −1.32 | $p$ = 0.188, NS |
| | Mixed | −0.0322 | 0.968 (CI 0.641 to 1.46) | 0.211 | −0.153 | $p$ = 0.879, NS |
| | Other | −0.770 | 0.463 (CI 0.255 to 0.839) | 0.303 | −2.54 | $p$ = 0.0111 * |
| | Unknown | −0.192 | 0.825 (CI 0.711 to 0.959) | 0.0763 | −2.51 | $p$ = 0.0119 * |

| Term | Level | Coefficient | $e^{coeff}$ | SE(coeff) | $Z$ | $p_{\|Z\|}$ |
|---|---|---|---|---|---|---|
| IMD fraction (0–1, higher values indicating less deprived) | – | −0.00196 | 0.998 (CI 0.834 to 1.19) | 0.0915 | −0.0214 | $p$ = 0.983, NS |
| Organic mental disorders (F0) | True | −0.370 | 0.691 (CI 0.579 to 0.824) | 0.0902 | −4.10 | $p = 4.11 \times 10^{-5}$ **** |
| Substance misuse (F1) | True | −0.0319 | 0.969 (CI 0.803 to 1.17) | 0.0954 | −0.334 | $p$ = 0.738, NS |
| Psychotic disorders (F2) | True | −0.327 | 0.721 (CI 0.562 to 0.926) | 0.127 | −2.56 | $p$ = 0.0104 * |
| Manic episode and bipolar disorder (F30–F31) | True | −0.383 | 0.682 (CI 0.566 to 0.821) | 0.0949 | −4.04 | $p = 5.45 \times 10^{-5}$ **** |
| Anxiety disorders and stress reactions (F40–F43) | True | −0.0815 | 0.922 (CI 0.834 to 1.02) | 0.0512 | −1.59 | $p$ = 0.112, NS |
| Eating disorders (F50) | True | −0.393 | 0.675 (CI 0.508 to 0.897) | 0.145 | −2.71 | $p$ = 0.00668 ** |
| Personality disorders (F60–F61) | True | +0.354 | 1.42 (CI 1.24 to 1.64) | 0.0715 | +4.95 | $p = 7.45 \times 10^{-7}$ ***** |
| Intellectual disability (F7) | True | −2.26 | 0.105 (CI 0.0261 to 0.421) | 0.710 | −3.18 | $p$ = 0.00148 ** |
| Developmental disorders (F8) | True | −0.239 | 0.787 (CI 0.550 to 1.13) | 0.183 | −1.31 | $p$ = 0.192, NS |
| Childhood behavioural and emotional disorders (F9) | True | −0.665 | 0.514 (CI 0.339 to 0.779) | 0.212 | −3.13 | $p$ = 0.00172 ** |
| Intentional self-harm (X60–X84) | True | +0.139 | 1.15 (CI 0.917 to 1.44) | 0.115 | +1.20 | $p$ = 0.228, NS |

### Supplementary Table 10

***Predictors of TCA use***

| Term | Level | Coefficient | $e^{coeff}$ | SE(coeff) | $Z$ | $p_{\|Z\|}$ |
|---|---|---|---|---|---|---|
| Service (age band) at referral | Adult | Reference | | | | |
| | CAMHS | −1.05 | 0.351 (CI 0.215 to 0.571) | 0.249 | −4.21 | $p = 2.59 \times 10^{-5}$ **** |
| | Older adult | +0.391 | 1.48 (CI 1.29 to 1.69) | 0.0696 | +5.61 | $p = 2.03 \times 10^{-8}$ ***** |
| Sex | Female | Reference | | | | |
| | Male | −0.253 | 0.776 (CI 0.684 to 0.881) | 0.0644 | −3.94 | $p = 8.30 \times 10^{-5}$ **** |
| Ethnicity | White | Reference | | | | |
| | Asian | −0.313 | 0.731 (CI 0.463 to 1.15) | 0.233 | −1.34 | $p$ = 0.179, NS |
| | Black | +0.429 | 1.54 (CI 0.865 to 2.73) | 0.293 | +1.46 | $p$ = 0.143, NS |
| | Mixed | −0.412 | 0.663 (CI 0.343 to 1.28) | 0.336 | −1.23 | $p$ = 0.220, NS |
| | Other | +0.0863 | 1.09 (CI 0.615 to 1.93) | 0.292 | +0.296 | $p$ = 0.767, NS |
| | Unknown | +0.0515 | 1.05 (CI 0.885 to 1.25) | 0.0884 | +0.583 | $p$ = 0.560, NS |
| IMD fraction (0–1, higher values indicating less deprived) | – | +0.0281 | 1.03 (CI 0.820 to 1.29) | 0.116 | +0.243 | $p$ = 0.808, NS |

| Term | Level | Coefficient | $e^{coeff}$ | SE(coeff) | $Z$ | $p_{\|Z\|}$ |
|---|---|---|---|---|---|---|
| Organic mental disorders (F0) | True | −0.256 | 0.774 (CI 0.645 to 0.929) | 0.0931 | −2.75 | $p$ = 0.00589 ** |
| Substance misuse (F1) | True | −0.385 | 0.680 (CI 0.507 to 0.913) | 0.150 | −2.57 | $p$ = 0.0103 * |
| Psychotic disorders (F2) | True | −0.415 | 0.660 (CI 0.463 to 0.942) | 0.181 | −2.29 | $p$ = 0.0219 * |
| Manic episode and bipolar disorder (F30–F31) | True | −0.436 | 0.647 (CI 0.504 to 0.829) | 0.127 | −3.43 | $p$ = 0.000598 *** |
| Anxiety disorders and stress reactions (F40–F43) | True | −0.0545 | 0.947 (CI 0.833 to 1.08) | 0.0652 | −0.837 | $p$ = 0.403, NS |
| Eating disorders (F50) | True | −0.0564 | 0.945 (CI 0.668 to 1.34) | 0.177 | −0.319 | $p$ = 0.750, NS |
| Personality disorders (F60–F61) | True | +0.227 | 1.25 (CI 1.03 to 1.53) | 0.101 | +2.24 | $p$ = 0.0248 * |
| Intellectual disability (F7) | True | −0.327 | 0.721 (CI 0.318 to 1.63) | 0.418 | −0.783 | $p$ = 0.434, NS |
| Developmental disorders (F8) | True | −0.309 | 0.734 (CI 0.436 to 1.23) | 0.265 | −1.17 | $p$ = 0.244, NS |
| Childhood behavioural and emotional disorders (F9) | True | −0.564 | 0.569 (CI 0.320 to 1.01) | 0.293 | −1.92 | $p$ = 0.0545, NS |
| Intentional self-harm (X60–X84) | True | −0.0495 | 0.952 (CI 0.693 to 1.31) | 0.162 | −0.306 | $p$ = 0.760, NS |

### Supplementary Table 11

***Predictors of lamotrigine use***

| Term | Level | Coefficient | $e^{coeff}$ | SE(coeff) | $Z$ | $p_{\|Z\|}$ |
|---|---|---|---|---|---|---|
| Service (age band) at referral | Adult | Reference | | | | |
| | CAMHS | −0.770 | 0.463 (CI 0.236 to 0.910) | 0.344 | −2.24 | $p$ = 0.0254 * |
| | Older adult | −1.61 | 0.201 (CI 0.137 to 0.294) | 0.195 | −8.26 | $p < 2.20 \times 10^{-16}$ ***** |
| Sex | Female | Reference | | | | |
| | Male | −0.102 | 0.903 (CI 0.719 to 1.13) | 0.117 | −0.874 | $p$ = 0.382, NS |
| Ethnicity | White | Reference | | | | |
| | Asian | −0.607 | 0.545 (CI 0.225 to 1.32) | 0.452 | −1.34 | $p$ = 0.180, NS |
| | Black | −0.674 | 0.510 (CI 0.162 to 1.60) | 0.584 | −1.15 | $p$ = 0.249, NS |
| | Mixed | +0.0954 | 1.10 (CI 0.453 to 2.67) | 0.453 | +0.211 | $p$ = 0.833, NS |
| | Other | −0.682 | 0.505 (CI 0.125 to 2.04) | 0.711 | −0.959 | $p$ = 0.337, NS |
| | Unknown | −0.223 | 0.800 (CI 0.541 to 1.18) | 0.200 | −1.11 | $p$ = 0.266, NS |
| IMD fraction (0–1, higher values indicating less deprived) | – | +0.0861 | 1.09 (CI 0.729 to 1.63) | 0.205 | +0.419 | $p$ = 0.675, NS |
| Organic mental disorders (F0) | True | +0.361 | 1.43 (CI 0.906 to 2.27) | 0.235 | +1.54 | $p$ = 0.124, NS |

| Term | Level | Coefficient | $e^{coeff}$ | SE(coeff) | $Z$ | $p_{\|Z\|}$ |
|---|---|---|---|---|---|---|
| Substance misuse (F1) | True | −0.0734 | 0.929 (CI 0.619 to 1.40) | 0.207 | −0.354 | $p$ = 0.724, NS |
| Psychotic disorders (F2) | True | −0.0898 | 0.914 (CI 0.560 to 1.49) | 0.250 | −0.359 | $p$ = 0.720, NS |
| Manic episode and bipolar disorder (F30–F31) | True | +2.04 | 7.71 (CI 6.19 to 9.61) | 0.112 | +18.2 | $p < 2.20 \times 10^{-16}$ ***** |
| Anxiety disorders and stress reactions (F40–F43) | True | −0.337 | 0.714 (CI 0.549 to 0.927) | 0.134 | −2.53 | $p$ = 0.0115 * |
| Eating disorders (F50) | True | +0.0794 | 1.08 (CI 0.647 to 1.81) | 0.263 | +0.302 | $p$ = 0.763, NS |
| Personality disorders (F60–F61) | True | +0.322 | 1.38 (CI 1.04 to 1.83) | 0.143 | +2.25 | $p$ = 0.0245 * |
| Intellectual disability (F7) | True | −0.0883 | 0.915 (CI 0.282 to 2.98) | 0.601 | −0.147 | $p$ = 0.883, NS |
| Developmental disorders (F8) | True | −0.585 | 0.557 (CI 0.224 to 1.39) | 0.466 | −1.26 | $p$ = 0.209, NS |
| Childhood behavioural and emotional disorders (F9) | True | −0.616 | 0.540 (CI 0.200 to 1.46) | 0.508 | −1.21 | $p$ = 0.225, NS |
| Intentional self-harm (X60–X84) | True | −0.241 | 0.786 (CI 0.439 to 1.41) | 0.298 | −0.810 | $p$ = 0.418, NS |

### Supplementary Table 12

***Predictors of lithium use***

| Term | Level | Coefficient | $e^{coeff}$ | SE(coeff) | $Z$ | $p_{\|Z\|}$ |
|---|---|---|---|---|---|---|
| Service (age band) at referral | Adult | Reference | | | | |
| | CAMHS | −1.22 | 0.296 (CI 0.121 to 0.722) | 0.456 | −2.68 | $p$ = 0.00745 ** |
| | Older adult | +0.225 | 1.25 (CI 1.01 to 1.55) | 0.107 | +2.09 | $p$ = 0.0366 * |
| Sex | Female | Reference | | | | |
| | Male | −0.0186 | 0.982 (CI 0.809 to 1.19) | 0.0988 | −0.188 | $p$ = 0.851, NS |
| Ethnicity | White | Reference | | | | |
| | Asian | −0.527 | 0.590 (CI 0.263 to 1.33) | 0.413 | −1.28 | $p$ = 0.201, NS |
| | Black | −0.0930 | 0.911 (CI 0.375 to 2.21) | 0.452 | −0.206 | $p$ = 0.837, NS |
| | Mixed | −0.528 | 0.590 (CI 0.189 to 1.84) | 0.581 | −0.910 | $p$ = 0.363, NS |
| | Other | −1.66 | 0.190 (CI 0.0266 to 1.35) | 1.00 | −1.66 | $p$ = 0.0972, NS |
| | Unknown | −0.532 | 0.587 (CI 0.413 to 0.835) | 0.180 | −2.96 | $p$ = 0.00308 ** |
| IMD fraction (0–1, higher values indicating less deprived) | – | +0.714 | 2.04 (CI 1.41 to 2.95) | 0.188 | +3.80 | $p$ = 0.000147 *** |
| Organic mental disorders (F0) | True | −0.210 | 0.811 (CI 0.596 to 1.10) | 0.157 | −1.33 | $p$ = 0.182, NS |
| Substance misuse (F1) | True | −0.286 | 0.752 (CI 0.491 to 1.15) | 0.217 | −1.32 | $p$ = 0.188, NS |

| Term | Level | Coefficient | $e^{coeff}$ | SE(coeff) | $Z$ | $p_{\|Z\|}$ |
|---|---|---|---|---|---|---|
| Psychotic disorders (F2) | True | −0.225 | 0.798 (CI 0.490 to 1.30) | 0.249 | −0.904 | $p$ = 0.366, NS |
| Manic episode and bipolar disorder (F30–F31) | True | +2.00 | 7.35 (CI 6.04 to 8.95) | 0.101 | +19.8 | $p < 2.20 \times 10^{-16}$ ***** |
| Anxiety disorders and stress reactions (F40–F43) | True | −0.519 | 0.595 (CI 0.467 to 0.757) | 0.123 | −4.22 | $p = 2.45 \times 10^{-5}$ **** |
| Eating disorders (F50) | True | −0.532 | 0.587 (CI 0.288 to 1.20) | 0.363 | −1.47 | $p$ = 0.142, NS |
| Personality disorders (F60–F61) | True | −0.0216 | 0.979 (CI 0.721 to 1.33) | 0.156 | −0.138 | $p$ = 0.890, NS |
| Intellectual disability (F7) | True | −0.164 | 0.848 (CI 0.206 to 3.49) | 0.721 | −0.228 | $p$ = 0.820, NS |
| Developmental disorders (F8) | True | −1.48 | 0.228 (CI 0.0558 to 0.935) | 0.719 | −2.05 | $p$ = 0.0400 * |
| Childhood behavioural and emotional disorders (F9) | True | −0.160 | 0.852 (CI 0.350 to 2.08) | 0.455 | −0.351 | $p$ = 0.725, NS |
| Intentional self-harm (X60–X84) | True | +0.171 | 1.19 (CI 0.743 to 1.89) | 0.239 | +0.715 | $p$ = 0.474, NS |

### Supplementary Table 13

***Predictors of trazodone use***

| Term | Level | Coefficient | $e^{coeff}$ | SE(coeff) | $Z$ | $p_{\|Z\|}$ |
|---|---|---|---|---|---|---|
| Service (age band) at referral | Adult | Reference | | | | |
| | CAMHS | −1.51 | 0.221 (CI 0.0537 to 0.909) | 0.722 | −2.09 | $p$ = 0.0364 * |
| | Older adult | −0.293 | 0.746 (CI 0.492 to 1.13) | 0.213 | −1.38 | $p$ = 0.169, NS |
| Sex | Female | Reference | | | | |
| | Male | +0.0474 | 1.05 (CI 0.749 to 1.47) | 0.171 | +0.277 | $p$ = 0.782, NS |
| Ethnicity | White | Reference | | | | |
| | Asian | −0.552 | 0.576 (CI 0.141 to 2.34) | 0.716 | −0.771 | $p$ = 0.441, NS |
| | Black | −0.317 | 0.728 (CI 0.101 to 5.26) | 1.01 | −0.314 | $p$ = 0.754, NS |
| | Mixed | +1.27 | 3.56 (CI 1.55 to 8.14) | 0.422 | +3.00 | $p$ = 0.00266 ** |
| | Other | +0.189 | 1.21 (CI 0.297 to 4.92) | 0.716 | +0.264 | $p$ = 0.792, NS |
| | Unknown | +0.0253 | 1.03 (CI 0.612 to 1.72) | 0.263 | +0.0960 | $p$ = 0.924, NS |
| IMD fraction (0–1, higher values indicating less deprived) | – | +0.270 | 1.31 (CI 0.710 to 2.42) | 0.312 | +0.863 | $p$ = 0.388, NS |
| Organic mental disorders (F0) | True | +0.201 | 1.22 (CI 0.711 to 2.10) | 0.276 | +0.726 | $p$ = 0.468, NS |
| Substance misuse (F1) | True | −0.376 | 0.687 (CI 0.333 to 1.42) | 0.369 | −1.02 | $p$ = 0.308, NS |
| Psychotic disorders (F2) | True | −0.0430 | 0.958 (CI 0.445 to 2.06) | 0.391 | −0.110 | $p$ = 0.912, NS |
| Manic episode and bipolar disorder (F30–F31) | True | −0.119 | 0.888 (CI 0.499 to 1.58) | 0.295 | −0.402 | $p$ = 0.687, NS |
| Anxiety disorders and stress reactions (F40–F43) | True | +0.00899 | 1.01 (CI 0.716 to 1.42) | 0.175 | +0.0515 | $p$ = 0.959, NS |

| Term | Level | Coefficient | $e^{coeff}$ | SE(coeff) | $Z$ | $p_{\|Z\|}$ |
|---|---|---|---|---|---|---|
| Eating disorders (F50) | True | +0.279 | 1.32 (CI 0.632 to 2.76) | 0.376 | +0.740 | $p$ = 0.459, NS |
| Personality disorders (F60–F61) | True | +0.775 | 2.17 (CI 1.42 to 3.31) | 0.216 | +3.59 | $p$ = 0.000326 *** |
| Intellectual disability (F7) | True | −0.125 | 0.883 (CI 0.120 to 6.50) | 1.02 | −0.123 | $p$ = 0.902, NS |
| Developmental disorders (F8) | True | −1.35 | 0.260 (CI 0.0356 to 1.90) | 1.01 | −1.33 | $p$ = 0.184, NS |
| Childhood behavioural and emotional disorders (F9) | True | −0.437 | 0.646 (CI 0.158 to 2.64) | 0.719 | −0.608 | $p$ = 0.543, NS |
| Intentional self-harm (X60–X84) | True | −0.00371 | 0.996 (CI 0.460 to 2.16) | 0.394 | −0.00942 | $p$ = 0.992, NS |

### Supplementary Table 14

***Predictors of MAOI use***

| Term | Level | Coefficient | $e^{coeff}$ | SE(coeff) | $Z$ | $p_{\|Z\|}$ |
|---|---|---|---|---|---|---|
| Service (age band) at referral | Adult | Reference | | | | |
| | CAMHS | −18.0 | $1.59 \times 10^{-8}$ (CI 0.00 to Inf) | $4.98 \times 10^{3}$ | −0.00361 | $p$ = 0.997, NS |
| | Older adult | +0.00635 | 1.01 (CI 0.518 to 1.95) | 0.338 | +0.0188 | $p$ = 0.985, NS |
| Sex | Female | Reference | | | | |
| | Male | +0.357 | 1.43 (CI 0.808 to 2.53) | 0.291 | +1.23 | $p$ = 0.219, NS |
| Ethnicity | White | Reference | | | | |
| | Asian | −18.2 | $1.24 \times 10^{-8}$ (CI 0.00 to Inf) | $8.03 \times 10^{3}$ | −0.00227 | $p$ = 0.998, NS |
| | Black | −18.3 | $1.17 \times 10^{-8}$ (CI 0.00 to Inf) | $1.27 \times 10^{4}$ | −0.00144 | $p$ = 0.999, NS |
| | Mixed | +0.612 | 1.84 (CI 0.251 to 13.5) | 1.02 | +0.602 | $p$ = 0.547, NS |
| | Other | −18.2 | $1.27 \times 10^{-8}$ (CI 0.00 to Inf) | $1.14 \times 10^{4}$ | −0.00160 | $p$ = 0.999, NS |
| | Unknown | −0.198 | 0.820 (CI 0.321 to 2.10) | 0.479 | −0.414 | $p$ = 0.679, NS |
| IMD fraction (0–1, higher values indicating less deprived) | – | +0.167 | 1.18 (CI 0.398 to 3.51) | 0.555 | +0.300 | $p$ = 0.764, NS |
| Organic mental disorders (F0) | True | −0.242 | 0.785 (CI 0.306 to 2.02) | 0.481 | −0.502 | $p$ = 0.616, NS |
| Substance misuse (F1) | True | +0.175 | 1.19 (CI 0.417 to 3.40) | 0.535 | +0.327 | $p$ = 0.744, NS |
| Psychotic disorders (F2) | True | +0.140 | 1.15 (CI 0.350 to 3.78) | 0.607 | +0.231 | $p$ = 0.817, NS |
| Manic episode and bipolar disorder (F30–F31) | True | −0.606 | 0.546 (CI 0.167 to 1.78) | 0.604 | −1.00 | $p$ = 0.316, NS |
| Anxiety disorders and stress reactions (F40–F43) | True | −0.262 | 0.769 (CI 0.410 to 1.44) | 0.322 | −0.816 | $p$ = 0.415, NS |
| Eating disorders (F50) | True | −17.7 | $1.98 \times 10^{-8}$ (CI 0.00 to Inf) | $5.80 \times 10^{3}$ | −0.00306 | $p$ = 0.998, NS |

| Term | Level | Coefficient | $e^{coeff}$ | SE(coeff) | $Z$ | $p_{\|Z\|}$ |
|---|---|---|---|---|---|---|
| Personality disorders (F60–F61) | True | −0.245 | 0.783 (CI 0.272 to 2.25) | 0.539 | −0.454 | $p$ = 0.650, NS |
| Intellectual disability (F7) | True | −18.2 | $1.29 \times 10^{-8}$ (CI 0.00 to Inf) | $1.10 \times 10^{4}$ | −0.00166 | $p$ = 0.999, NS |
| Developmental disorders (F8) | True | −0.260 | 0.771 (CI 0.102 to 5.84) | 1.03 | −0.252 | $p$ = 0.801, NS |
| Childhood behavioural and emotional disorders (F9) | True | +0.749 | 2.12 (CI 0.492 to 9.10) | 0.744 | +1.01 | $p$ = 0.314, NS |
| Intentional self-harm (X60–X84) | True | −0.758 | 0.469 (CI 0.0635 to 3.46) | 1.02 | −0.743 | $p$ = 0.458, NS |

### Supplementary Table 15

***Predictors of flupentixol use***

| Term | Level | Coefficient | $e^{coeff}$ | SE(coeff) | $Z$ | $p_{\|Z\|}$ |
|---|---|---|---|---|---|---|
| Service (age band) at referral | Adult | Reference | | | | |
| | CAMHS | −0.653 | 0.521 (CI 0.0706 to 3.84) | 1.02 | −0.640 | $p$ = 0.522, NS |
| | Older adult | −0.280 | 0.756 (CI 0.395 to 1.45) | 0.331 | −0.846 | $p$ = 0.398, NS |
| Sex | Female | Reference | | | | |
| | Male | +0.167 | 1.18 (CI 0.716 to 1.95) | 0.255 | +0.652 | $p$ = 0.514, NS |
| Ethnicity | White | Reference | | | | |
| | Asian | +0.578 | 1.78 (CI 0.546 to 5.82) | 0.604 | +0.958 | $p$ = 0.338, NS |
| | Black | +0.667 | 1.95 (CI 0.461 to 8.23) | 0.736 | +0.906 | $p$ = 0.365, NS |
| | Mixed | −17.7 | $1.99 \times 10^{-8}$ (CI 0.00 to Inf) | $7.59 \times 10^{3}$ | −0.00234 | $p$ = 0.998, NS |
| | Other | −17.8 | $1.83 \times 10^{-8}$ (CI 0.00 to Inf) | $6.85 \times 10^{3}$ | −0.00260 | $p$ = 0.998, NS |
| | Unknown | −17.7 | $1.97 \times 10^{-8}$ (CI 0.00 to Inf) | $2.68 \times 10^{3}$ | −0.00663 | $p$ = 0.995, NS |
| IMD fraction (0–1, higher values indicating less deprived) | – | +0.136 | 1.15 (CI 0.457 to 2.87) | 0.469 | +0.290 | $p$ = 0.772, NS |
| Organic mental disorders (F0) | True | −0.272 | 0.762 (CI 0.299 to 1.94) | 0.477 | −0.571 | $p$ = 0.568, NS |
| Substance misuse (F1) | True | +0.578 | 1.78 (CI 0.888 to 3.58) | 0.356 | +1.63 | $p$ = 0.104, NS |
| Psychotic disorders (F2) | True | +1.80 | 6.06 (CI 3.47 to 10.6) | 0.284 | +6.35 | $p = 2.14 \times 10^{-10}$ ***** |
| Manic episode and bipolar disorder (F30–F31) | True | +0.343 | 1.41 (CI 0.707 to 2.81) | 0.352 | +0.974 | $p$ = 0.330, NS |
| Anxiety disorders and stress reactions (F40–F43) | True | −0.684 | 0.505 (CI 0.272 to 0.937) | 0.316 | −2.17 | $p$ = 0.0303 * |
| Eating disorders (F50) | True | −1.08 | 0.340 (CI 0.0456 to 2.53) | 1.02 | −1.05 | $p$ = 0.292, NS |
| Personality disorders (F60–F61) | True | +0.771 | 2.16 (CI 1.16 to 4.03) | 0.318 | +2.42 | $p$ = 0.0154 * |

| Term | Level | Coefficient | $e^{coeff}$ | SE(coeff) | $Z$ | $p_{\|Z\|}$ |
|---|---|---|---|---|---|---|
| Intellectual disability (F7) | True | −17.5 | 2.46 × $10^{-8}$ (CI 0.00 to Inf) | 7.03 × $10^{3}$ | −0.00249 | $p$ = 0.998, NS |
| Developmental disorders (F8) | True | −0.103 | 0.902 (CI 0.123 to 6.60) | 1.02 | −0.102 | $p$ = 0.919, NS |
| Childhood behavioural and emotional disorders (F9) | True | −17.4 | 2.76 × $10^{-8}$ (CI 0.00 to Inf) | 5.26 × $10^{3}$ | −0.00331 | $p$ = 0.997, NS |
| Intentional self-harm (X60–X84) | True | −0.570 | 0.566 (CI 0.137 to 2.34) | 0.725 | −0.786 | $p$ = 0.432, NS |

### Supplementary Table 16

***Predictors of triiodothyronine use***

| Term | Level | Coefficient | $e^{coeff}$ | SE(coeff) | $Z$ | $p_{\|Z\|}$ |
|---|---|---|---|---|---|---|
| Service (age band) at referral | Adult | Reference | | | | |
| | CAMHS | +2.18 | 8.84 (CI 1.67 to 46.6) | 0.849 | +2.57 | $p$ = 0.0102 * |
| | Older adult | −23.4 | 6.90 × $10^{-11}$ (CI 0.00 to Inf) | 6.68 × $10^{4}$ | −0.000350 | $p$ = 1.00, NS |
| Sex | Female | Reference | | | | |
| | Male | −0.263 | 0.769 (CI 0.177 to 3.34) | 0.750 | −0.350 | $p$ = 0.726, NS |
| Ethnicity | White | Reference | | | | |
| | Asian | −17.9 | 1.69 × $10^{-8}$ (CI 0.00 to Inf) | 1.78 × $10^{4}$ | −0.00101 | $p$ = 0.999, NS |
| | Black | −19.0 | 5.66 × $10^{-9}$ (CI 0.00 to Inf) | 4.31 × $10^{4}$ | −0.000441 | $p$ = 1.00, NS |
| | Mixed | −17.5 | 2.59 × $10^{-8}$ (CI 0.00 to Inf) | 1.59 × $10^{4}$ | −0.00110 | $p$ = 0.999, NS |
| | Other | −18.3 | 1.10 × $10^{-8}$ (CI 0.00 to Inf) | 1.88 × $10^{4}$ | −0.000976 | $p$ = 0.999, NS |
| | Unknown | −0.255 | 0.775 (CI 0.0931 to 6.45) | 1.08 | −0.236 | $p$ = 0.814, NS |
| IMD fraction (0–1, higher values indicating less deprived) | – | +3.30 | 27.0 (CI 0.834 to 874) | 1.77 | +1.86 | $p$ = 0.0632, NS |
| Organic mental disorders (F0) | True | −16.6 | 6.49 × $10^{-8}$ (CI 0.00 to Inf) | 1.43 × $10^{4}$ | −0.00116 | $p$ = 0.999, NS |
| Substance misuse (F1) | True | +1.80 | 6.08 (CI 1.15 to 32.2) | 0.851 | +2.12 | $p$ = 0.0339 * |
| Psychotic disorders (F2) | True | +1.06 | 2.90 (CI 0.331 to 25.4) | 1.11 | +0.961 | $p$ = 0.337, NS |
| Manic episode and bipolar disorder (F30–F31) | True | −18.9 | 6.45 × $10^{-9}$ (CI 0.00 to Inf) | 1.04 × $10^{4}$ | −0.00182 | $p$ = 0.999, NS |
| Anxiety disorders and stress reactions (F40–F43) | True | −16.9 | 4.81 × $10^{-8}$ (CI 0.00 to Inf) | 2.11 × $10^{3}$ | −0.00798 | $p$ = 0.994, NS |
| Eating disorders (F50) | True | −18.9 | 6.33 × $10^{-9}$ (CI 0.00 to Inf) | 1.14 × $10^{4}$ | −0.00166 | $p$ = 0.999, NS |
| Personality disorders (F60–F61) | True | −17.3 | 3.06 × $10^{-8}$ (CI 0.00 to Inf) | 5.66 × $10^{3}$ | −0.00305 | $p$ = 0.998, NS |
| Intellectual disability (F7) | True | −16.3 | 8.16 × $10^{-8}$ (CI 0.00 to Inf) | 1.98 × $10^{4}$ | −0.000823 | $p$ = 0.999, NS |
| Developmental disorders (F8) | True | −16.6 | 6.04 × $10^{-8}$ (CI 0.00 to Inf) | 1.04 × $10^{4}$ | −0.00159 | $p$ = 0.999, NS |

| Term | Level | Coefficient | $e^{coeff}$ | SE(coeff) | $Z$ | $p_{\|Z\|}$ |
|---|---|---|---|---|---|---|
| Childhood behavioural and emotional disorders (F9) | True | −18.4 | $9.98 \times 10^{-9}$ (CI 0.00 to Inf) | $1.61 \times 10^{4}$ | −0.00114 | $p = 0.999$, NS |
| Intentional self-harm (X60–X84) | True | −14.5 | $5.21 \times 10^{-7}$ (CI 0.00 to Inf) | $1.98 \times 10^{3}$ | −0.00731 | $p = 0.994$, NS |

### Supplementary Table 17

***Predictors of psychology input***

| Term | Level | Coefficient | $e^{coeff}$ | SE(coeff) | $Z$ | $p_{\|Z\|}$ |
|---|---|---|---|---|---|---|
| Service (age band) at referral | Adult | Reference | | | | |
| | CAMHS | +0.657 | 1.93 (CI 1.65 to 2.25) | 0.0793 | +8.28 | $p < 2.20 \times 10^{-16}$ ***** |
| | Older adult | +0.412 | 1.51 (CI 1.38 to 1.64) | 0.0438 | +9.39 | $p < 2.20 \times 10^{-16}$ ***** |
| Sex | Female | Reference | | | | |
| | Male | +0.0558 | 1.06 (CI 0.980 to 1.14) | 0.0388 | +1.44 | $p = 0.151$, NS |
| Ethnicity | White | Reference | | | | |
| | Asian | −0.0200 | 0.980 (CI 0.769 to 1.25) | 0.124 | −0.161 | $p = 0.872$, NS |
| | Black | −0.270 | 0.764 (CI 0.480 to 1.22) | 0.237 | −1.14 | $p = 0.256$, NS |
| | Mixed | −0.0236 | 0.977 (CI 0.713 to 1.34) | 0.160 | −0.147 | $p = 0.883$, NS |
| | Other | +0.189 | 1.21 (CI 0.872 to 1.67) | 0.166 | +1.14 | $p = 0.256$, NS |
| | Unknown | −0.452 | 0.636 (CI 0.559 to 0.723) | 0.0655 | −6.91 | $p = 4.91 \times 10^{-12}$ ***** |
| IMD fraction (0–1, higher values indicating less deprived) | – | +0.0375 | 1.04 (CI 0.903 to 1.19) | 0.0709 | +0.528 | $p = 0.597$, NS |
| Organic mental disorders (F0) | True | −0.737 | 0.479 (CI 0.419 to 0.547) | 0.0679 | −10.9 | $p < 2.20 \times 10^{-16}$ ***** |
| Substance misuse (F1) | True | −0.157 | 0.855 (CI 0.731 to 1.00) | 0.0799 | −1.96 | $p = 0.0496$ * |
| Psychotic disorders (F2) | True | −0.113 | 0.893 (CI 0.746 to 1.07) | 0.0919 | −1.23 | $p = 0.217$, NS |
| Manic episode and bipolar disorder (F30–F31) | True | −0.170 | 0.844 (CI 0.738 to 0.965) | 0.0684 | −2.49 | $p = 0.0129$ * |
| Anxiety disorders and stress reactions (F40–F43) | True | +0.398 | 1.49 (CI 1.38 to 1.60) | 0.0381 | +10.4 | $p < 2.20 \times 10^{-16}$ ***** |
| Eating disorders (F50) | True | +0.993 | 2.70 (CI 2.31 to 3.16) | 0.0802 | +12.4 | $p < 2.20 \times 10^{-16}$ ***** |
| Personality disorders (F60–F61) | True | +0.180 | 1.20 (CI 1.07 to 1.34) | 0.0577 | +3.12 | $p = 0.00182$ ** |
| Intellectual disability (F7) | True | −0.756 | 0.469 (CI 0.285 to 0.773) | 0.254 | −2.97 | $p = 0.00295$ ** |
| Developmental disorders (F8) | True | +0.209 | 1.23 (CI 0.979 to 1.55) | 0.118 | +1.78 | $p = 0.0758$, NS |
| Childhood behavioural and emotional disorders (F9) | True | −0.473 | 0.623 (CI 0.482 to 0.806) | 0.131 | −3.61 | $p = 0.000312$ *** |

| Term | Level | Coefficient | $e^{coeff}$ | SE(coeff) | $Z$ | $p_{\|Z\|}$ |
|---|---|---|---|---|---|---|
| Intentional self-harm (X60–X84) | True | −0.134 | 0.875 (CI 0.724 to 1.06) | 0.0962 | −1.39 | $p$ = 0.163, NS |

**Supplementary Table 18**

*Predictors of crisis team referral*

| Term | Level | Coefficient | $e^{coeff}$ | SE(coeff) | $Z$ | $p_{\|Z\|}$ |
|---|---|---|---|---|---|---|
| Service (age band) at referral | Adult | Reference | | | | |
| | CAMHS | −2.01 | 0.134 (CI 0.0695 to 0.260) | 0.337 | −5.96 | $p = 2.50 \times 10^{-9}$ ***** |
| | Older adult | +0.0252 | 1.03 (CI 0.899 to 1.17) | 0.0670 | +0.376 | $p$ = 0.707, NS |
| Sex | Female | Reference | | | | |
| | Male | +0.150 | 1.16 (CI 1.04 to 1.30) | 0.0560 | +2.68 | $p$ = 0.00735 ** |
| Ethnicity | White | Reference | | | | |
| | Asian | +0.269 | 1.31 (CI 0.959 to 1.79) | 0.159 | +1.70 | $p$ = 0.0901, NS |
| | Black | +0.618 | 1.85 (CI 1.18 to 2.93) | 0.233 | +2.65 | $p$ = 0.00795 ** |
| | Mixed | −0.384 | 0.681 (CI 0.385 to 1.20) | 0.291 | −1.32 | $p$ = 0.186, NS |
| | Other | −0.00254 | 0.997 (CI 0.608 to 1.64) | 0.253 | −0.0100 | $p$ = 0.992, NS |
| | Unknown | −0.684 | 0.504 (CI 0.409 to 0.622) | 0.107 | −6.39 | $p = 1.66 \times 10^{-10}$ ***** |
| IMD fraction (0–1, higher values indicating less deprived) | – | −0.0835 | 0.920 (CI 0.749 to 1.13) | 0.105 | −0.795 | $p$ = 0.427, NS |
| Organic mental disorders (F0) | True | −0.169 | 0.844 (CI 0.704 to 1.01) | 0.0928 | −1.82 | $p$ = 0.0681, NS |
| Substance misuse (F1) | True | +0.477 | 1.61 (CI 1.36 to 1.92) | 0.0882 | +5.41 | $p = 6.36 \times 10^{-8}$ ***** |
| Psychotic disorders (F2) | True | +0.572 | 1.77 (CI 1.44 to 2.18) | 0.106 | +5.38 | $p = 7.44 \times 10^{-8}$ ***** |
| Manic episode and bipolar disorder (F30–F31) | True | +0.209 | 1.23 (CI 1.03 to 1.47) | 0.0913 | +2.29 | $p$ = 0.0219 * |
| Anxiety disorders and stress reactions (F40–F43) | True | −0.127 | 0.881 (CI 0.784 to 0.991) | 0.0598 | −2.12 | $p$ = 0.0343 * |
| Eating disorders (F50) | True | −0.312 | 0.732 (CI 0.522 to 1.03) | 0.173 | −1.81 | $p$ = 0.0703, NS |
| Personality disorders (F60–F61) | True | +0.526 | 1.69 (CI 1.45 to 1.97) | 0.0780 | +6.75 | $p = 1.47 \times 10^{-11}$ ***** |
| Intellectual disability (F7) | True | −1.44 | 0.236 (CI 0.0754 to 0.739) | 0.582 | −2.48 | $p$ = 0.0131 * |
| Developmental disorders (F8) | True | −0.0957 | 0.909 (CI 0.603 to 1.37) | 0.209 | −0.458 | $p$ = 0.647, NS |
| Childhood behavioural and emotional disorders (F9) | True | −0.701 | 0.496 (CI 0.302 to 0.817) | 0.254 | −2.76 | $p$ = 0.00584 ** |
| Intentional self-harm (X60–X84) | True | +1.26 | 3.52 (CI 2.94 to 4.20) | 0.0909 | +13.8 | $p < 2.20 \times 10^{-16}$ ***** |

### Supplementary Table 19

***Predictors of inpatient admission***

| Term | Level | Coefficient | $e^{coeff}$ | SE(coeff) | $Z$ | $p_{\|Z\|}$ |
|---|---|---|---|---|---|---|
| Service (age band) at referral | Adult | Reference | | | | |
| | CAMHS | +0.721 | 2.06 (CI 1.65 to 2.57) | 0.113 | +6.39 | $p = 1.71 \times 10^{-10}$ ***** |
| | Older adult | −0.274 | 0.761 (CI 0.652 to 0.888) | 0.0789 | −3.47 | $p$ = 0.000518 *** |
| Sex | Female | Reference | | | | |
| | Male | +0.200 | 1.22 (CI 1.08 to 1.38) | 0.0622 | +3.21 | $p$ = 0.00132 ** |
| Ethnicity | White | Reference | | | | |
| | Asian | −0.289 | 0.749 (CI 0.484 to 1.16) | 0.222 | −1.30 | $p$ = 0.193, NS |
| | Black | +0.234 | 1.26 (CI 0.714 to 2.24) | 0.292 | +0.804 | $p$ = 0.422, NS |
| | Mixed | −0.702 | 0.496 (CI 0.257 to 0.957) | 0.336 | −2.09 | $p$ = 0.0365 * |
| | Other | −0.301 | 0.740 (CI 0.408 to 1.34) | 0.304 | −0.988 | $p$ = 0.323, NS |
| | Unknown | −0.623 | 0.536 (CI 0.427 to 0.673) | 0.116 | −5.38 | $p = 7.49 \times 10^{-8}$ ***** |
| IMD fraction (0–1, higher values indicating less deprived) | – | −0.383 | 0.682 (CI 0.546 to 0.851) | 0.113 | −3.38 | $p$ = 0.000726 *** |
| Organic mental disorders (F0) | True | −0.0854 | 0.918 (CI 0.740 to 1.14) | 0.110 | −0.776 | $p$ = 0.438, NS |
| Substance misuse (F1) | True | +0.817 | 2.26 (CI 1.90 to 2.69) | 0.0886 | +9.22 | $p < 2.20 \times 10^{-16}$ ***** |
| Psychotic disorders (F2) | True | +0.646 | 1.91 (CI 1.53 to 2.38) | 0.113 | +5.71 | $p = 1.10 \times 10^{-8}$ ***** |
| Manic episode and bipolar disorder (F30–F31) | True | +0.281 | 1.33 (CI 1.09 to 1.61) | 0.0994 | +2.83 | $p$ = 0.00464 ** |
| Anxiety disorders and stress reactions (F40–F43) | True | −0.109 | 0.897 (CI 0.791 to 1.02) | 0.0643 | −1.69 | $p$ = 0.0907, NS |
| Eating disorders (F50) | True | +0.856 | 2.35 (CI 1.89 to 2.94) | 0.113 | +7.60 | $p = 2.93 \times 10^{-14}$ ***** |
| Personality disorders (F60–F61) | True | +0.366 | 1.44 (CI 1.23 to 1.69) | 0.0819 | +4.47 | $p = 7.79 \times 10^{-6}$ ***** |
| Intellectual disability (F7) | True | −0.534 | 0.586 (CI 0.260 to 1.32) | 0.414 | −1.29 | $p$ = 0.198, NS |
| Developmental disorders (F8) | True | −0.254 | 0.776 (CI 0.510 to 1.18) | 0.214 | −1.18 | $p$ = 0.236, NS |
| Childhood behavioural and emotional disorders (F9) | True | −0.0204 | 0.980 (CI 0.690 to 1.39) | 0.179 | −0.114 | $p$ = 0.909, NS |
| Intentional self-harm (X60–X84) | True | +0.991 | 2.69 (CI 2.22 to 3.27) | 0.0994 | +9.97 | $p < 2.20 \times 10^{-16}$ ***** |

### Supplementary Table 20

***Predictors of ECT***

| Term | Level | Coefficient | $e^{coeff}$ | SE(coeff) | $Z$ | $p_{\|Z\|}$ |
|---|---|---|---|---|---|---|
| Service (age band) at referral | Adult | Reference | | | | |

| Term | Level | Coefficient | $e^{coeff}$ | SE(coeff) | $Z$ | $p_{\|Z\|}$ |
|---|---|---|---|---|---|---|
| | CAMHS | −17.8 | 1.92 × $10^{-8}$ (CI 0.00 to Inf) | 4.33 × $10^{3}$ | −0.00410 | $p$ = 0.997, NS |
| | Older adult | +0.176 | 1.19 (CI 0.677 to 2.10) | 0.288 | +0.609 | $p$ = 0.542, NS |
| Sex | Female | Reference | | | | |
| | Male | +0.303 | 1.35 (CI 0.820 to 2.23) | 0.256 | +1.18 | $p$ = 0.236, NS |
| Ethnicity | White | Reference | | | | |
| | Asian | +0.320 | 1.38 (CI 0.332 to 5.72) | 0.726 | +0.440 | $p$ = 0.660, NS |
| | Black | −18.0 | 1.46 × $10^{-8}$ (CI 0.00 to Inf) | 1.04 × $10^{4}$ | −0.00174 | $p$ = 0.999, NS |
| | Mixed | −18.0 | 1.59 × $10^{-8}$ (CI 0.00 to Inf) | 9.15 × $10^{3}$ | −0.00196 | $p$ = 0.998, NS |
| | Other | −18.2 | 1.26 × $10^{-8}$ (CI 0.00 to Inf) | 1.01 × $10^{4}$ | −0.00180 | $p$ = 0.999, NS |
| | Unknown | −2.06 | 0.127 (CI 0.0175 to 0.921) | 1.01 | −2.04 | $p$ = 0.0412 * |
| IMD fraction (0–1, higher values indicating less deprived) | – | +0.827 | 2.29 (CI 0.869 to 6.01) | 0.493 | +1.68 | $p$ = 0.0939, NS |
| Organic mental disorders (F0) | True | −0.148 | 0.862 (CI 0.398 to 1.87) | 0.395 | −0.375 | $p$ = 0.707, NS |
| Substance misuse (F1) | True | −0.544 | 0.581 (CI 0.179 to 1.88) | 0.600 | −0.906 | $p$ = 0.365, NS |
| Psychotic disorders (F2) | True | +0.0534 | 1.05 (CI 0.378 to 2.94) | 0.524 | +0.102 | $p$ = 0.919, NS |
| Manic episode and bipolar disorder (F30–F31) | True | −0.0947 | 0.910 (CI 0.408 to 2.03) | 0.410 | −0.231 | $p$ = 0.817, NS |
| Anxiety disorders and stress reactions (F40–F43) | True | −0.278 | 0.757 (CI 0.435 to 1.32) | 0.283 | −0.984 | $p$ = 0.325, NS |
| Eating disorders (F50) | True | −0.185 | 0.831 (CI 0.196 to 3.53) | 0.738 | −0.250 | $p$ = 0.803, NS |
| Personality disorders (F60–F61) | True | +0.415 | 1.52 (CI 0.733 to 3.13) | 0.370 | +1.12 | $p$ = 0.262, NS |
| Intellectual disability (F7) | True | −17.9 | 1.75 × $10^{-8}$ (CI 0.00 to Inf) | 7.83 × $10^{3}$ | −0.00228 | $p$ = 0.998, NS |
| Developmental disorders (F8) | True | −0.176 | 0.838 (CI 0.115 to 6.13) | 1.02 | −0.174 | $p$ = 0.862, NS |
| Childhood behavioural and emotional disorders (F9) | True | −17.6 | 2.28 × $10^{-8}$ (CI 0.00 to Inf) | 5.84 × $10^{3}$ | −0.00301 | $p$ = 0.998, NS |
| Intentional self-harm (X60–X84) | True | +0.337 | 1.40 (CI 0.497 to 3.95) | 0.529 | +0.637 | $p$ = 0.524, NS |

### Supplementary Table 21

***Predictors of change in HoNOS scores (last minus first, more negative changes are better), for those with two such scores at least 7 days apart. EXCLUDES those with a lifetime diagnosis of bipolar affective disorder.***

| Term | $F$ (via Type III sums of squares) | $p_F$ | Level | Coefficient (95% CI) | Standard error | $t$ | $p_t$ |
|---|---|---|---|---|---|---|---|
| **DEPRESSION SUB-SCORES** | | | | | | | |
| (Intercept) | | | – | −0.628 (CI −0.780 to −0.476) | 0.0775 | $t_{2933}$ = −8.09 | $p$ = 8.41 × $10^{-16}$ ***** |
| Service (age band) at referral | $F_{2,2933}$ = 12.1 | $p$ = 5.79 × $10^{-6}$ ***** | Adult | Reference | | | |

| Term | $F$ (via Type III sums of squares) | $p_F$ | Level | Coefficient (95% CI) | Standard error | $t$ | $p_t$ |
|---|---|---|---|---|---|---|---|
| | | | CAMHS | +0.436 (CI −0.0367 to +0.909) | 0.241 | $t_{2933}$ = +1.81 | $p$ = 0.0706, NS |
| | | | Older adult | −0.237 (CI −0.341 to −0.133) | 0.0530 | $t_{2933}$ = −4.47 | $p = 8.14 \times 10^{-6}$ ***** |
| Sex | $F < 1$ | NS | | | | | |
| Ethnicity | $F < 1$ | NS | | | | | |
| IMD fraction (0–1, higher values indicating less deprived) | $F_{1,2933} = 4.08$ | $p$ = 0.0436 * | – | −0.163 (CI −0.322 to −0.00469) | 0.0809 | $t_{2933}$ = −2.02 | $p$ = 0.0436 * |
| Organic mental disorders (F0) | $F_{1,2933} = 27.1$ | $p = 2.09 \times 10^{-7}$ ***** | True | +0.334 (CI +0.208 to +0.460) | 0.0642 | $t_{2933}$ = +5.20 | $p = 2.09 \times 10^{-7}$ ***** |
| Substance misuse (F1) | $F_{1,2933} = 12.5$ | $p$ = 0.000415 *** | True | +0.311 (CI +0.139 to +0.484) | 0.0880 | $t_{2933}$ = +3.53 | $p$ = 0.000415 *** |
| Psychotic disorders (F2) | $F < 1$ | NS | | | | | |
| Anxiety disorders and stress reactions (F40–F43) | $F_{1,2933} = 7.15$ | $p$ = 0.00753 ** | True | +0.119 (CI +0.0317 to +0.206) | 0.0444 | $t_{2933}$ = +2.67 | $p$ = 0.00753 ** |
| Eating disorders (F50) | $F_{1,2933} = 3.14$ | $p$ = 0.0765, NS | | | | | |
| Personality disorders (F60–F61) | $F_{1,2933} = 38.0$ | $p = 8.17 \times 10^{-10}$ ***** | True | +0.446 (CI +0.304 to +0.587) | 0.0723 | $t_{2933}$ = +6.16 | $p = 8.17 \times 10^{-10}$ ***** |
| Intellectual disability (F7) | $F_{1,2933} = 1.30$ | $p$ = 0.255, NS | | | | | |
| Developmental disorders (F8) | $F < 1$ | NS | | | | | |
| Childhood behavioural and emotional disorders (F9) | $F_{1,2933} = 3.10$ | $p$ = 0.0784, NS | | | | | |
| Intentional self-harm (X60–X84) | $F_{1,2933} = 3.69$ | $p$ = 0.0547, NS | | | | | |
| Selective serotonin reuptake inhibitor (SSRI) | $F_{1,2933} = 12.8$ | $p$ = 0.000356 *** | True | −0.163 (CI −0.253 to −0.0736) | 0.0456 | $t_{2933}$ = −3.57 | $p$ = 0.000356 *** |
| Tricyclic antidepressant (TCA) | $F < 1$ | NS | | | | | |
| Monoamine oxidase inhibitor (MAOI) | $F < 1$ | NS | | | | | |
| Serotonin/noradrenaline reuptake inhibitor (SNRI) | $F < 1$ | NS | | | | | |
| Mirtazapine | $F < 1$ | NS | | | | | |
| Trazodone | $F < 1$ | NS | | | | | |
| Antipsychotics endorsed by NICE for depression | $F_{1,2933} = 1.07$ | $p$ = 0.302, NS | | | | | |
| Flupentixol | $F < 1$ | NS | | | | | |
| Lithium | $F_{1,2933} = 3.18$ | $p$ = 0.0746, NS | | | | | |
| Lamotrigine | $F < 1$ | NS | | | | | |
| Electroconvulsive therapy (ECT) | $F < 1$ | NS | | | | | |
| Psychology input | $F_{1,2933} = 3.68$ | $p$ = 0.0551, NS | | | | | |

**TOTAL SCORES**

| Term | $F$ (via Type III sums of squares) | $p_F$ | Level | Coefficient (95% CI) | Standard error | $t$ | $p_t$ |
|---|---|---|---|---|---|---|---|
| (Intercept) | | | – | −2.95 (CI −3.67 to −2.23) | 0.369 | $t_{2933}$ = −7.99 | $p$ = 1.85 × $10^{-15}$ ***** |
| Service (age band) at referral | $F$ < 1 | NS | | | | | |
| Sex | $F$ < 1 | NS | | | | | |
| Ethnicity | $F_{5,2933}$ = 1.17 | $p$ = 0.322, NS | | | | | |
| IMD fraction (0–1, higher values indicating less deprived) | $F_{1,2933}$ = 1.92 | $p$ = 0.166, NS | | | | | |
| Organic mental disorders (F0) | $F_{1,2933}$ = 34.3 | $p$ = 5.22 × $10^{-9}$ ***** | True | +1.79 (CI +1.19 to +2.39) | 0.306 | $t_{2933}$ = +5.86 | $p$ = 5.22 × $10^{-9}$ ***** |
| Substance misuse (F1) | $F_{1,2933}$ = 10.2 | $p$ = 0.00146 ** | True | +1.34 (CI +0.513 to +2.16) | 0.419 | $t_{2933}$ = +3.19 | $p$ = 0.00146 ** |
| Psychotic disorders (F2) | $F_{1,2933}$ = 7.96 | $p$ = 0.00482 ** | True | −1.35 (CI −2.29 to −0.411) | 0.478 | $t_{2933}$ = −2.82 | $p$ = 0.00482 ** |
| Anxiety disorders and stress reactions (F40–F43) | $F$ < 1 | NS | | | | | |
| Eating disorders (F50) | $F$ < 1 | NS | | | | | |
| Personality disorders (F60–F61) | $F_{1,2933}$ = 72.9 | $p$ < 2.20 × $10^{-16}$ ***** | True | +2.94 (CI +2.26 to +3.61) | 0.344 | $t_{2933}$ = +8.54 | $p$ < 2.20 × $10^{-16}$ ***** |
| Intellectual disability (F7) | $F_{1,2933}$ = 6.66 | $p$ = 0.00989 ** | True | +8.11 (CI +1.95 to +14.3) | 3.14 | $t_{2933}$ = +2.58 | $p$ = 0.00989 ** |
| Developmental disorders (F8) | $F$ < 1 | NS | | | | | |
| Childhood behavioural and emotional disorders (F9) | $F$ < 1 | NS | | | | | |
| Intentional self-harm (X60–X84) | $F_{1,2933}$ = 2.96 | $p$ = 0.0855, NS | | | | | |
| Selective serotonin reuptake inhibitor (SSRI) | $F_{1,2933}$ = 1.73 | $p$ = 0.189, NS | | | | | |
| Tricyclic antidepressant (TCA) | $F_{1,2933}$ = 5.62 | $p$ = 0.0178 * | True | +0.636 (CI +0.110 to +1.16) | 0.268 | $t_{2933}$ = +2.37 | $p$ = 0.0178 * |
| Monoamine oxidase inhibitor (MAOI) | $F_{1,2933}$ = 2.38 | $p$ = 0.123, NS | | | | | |
| Serotonin/noradrenaline reuptake inhibitor (SNRI) | $F$ < 1 | NS | | | | | |
| Mirtazapine | $F$ < 1 | NS | | | | | |
| Trazodone | $F$ < 1 | NS | | | | | |
| Antipsychotics endorsed by NICE for depression | $F_{1,2933}$ = 11.4 | $p$ = 0.000743 *** | True | −0.783 (CI −1.24 to −0.328) | 0.232 | $t_{2933}$ = −3.38 | $p$ = 0.000743 *** |
| Flupentixol | $F$ < 1 | NS | | | | | |
| Lithium | $F$ < 1 | NS | | | | | |
| Lamotrigine | $F_{1,2933}$ = 6.04 | $p$ = 0.0140 * | True | +1.33 (CI +0.269 to +2.39) | 0.540 | $t_{2933}$ = +2.46 | $p$ = 0.0140 * |
| Electroconvulsive therapy (ECT) | $F_{1,2933}$ = 1.50 | $p$ = 0.220, NS | | | | | |
| Psychology input | $F$ < 1 | NS | | | | | |
